# A canary in the mind: A single baseline brain scan predicts adolescent depression and anxiety one year later

**DOI:** 10.64898/2026.06.08.26355206

**Authors:** Gustavo Deco, Yonatan Sanz Perl, Jakub Vohryzek, Elvira Garcia-Guzman, Natasha Greenstein, Diego A. Pizzagalli, Ruben Laukkonen, Shamil Chandaria, Morten L. Kringelbach

## Abstract

Mood and anxiety disorders emerge predominantly in adolescence, yet they are usually identified only once symptoms have consolidated, when intervention can only be reactive. A marker that registers the loss of healthy brain function before symptoms crystallise would allow earlier and more targeted treatment, much as caged canaries once warned miners of danger before it became apparent. Here we report such a marker using a single baseline resting-state functional MRI scan in 150 adolescents in the Human Connectome Project Boston Adolescent Neuroimaging of Depression and Anxiety (HCP BANDA) cohort, allowing us to prospectively predict depression and anxiety symptoms one year later in held-out participants at r = 0.60, substantially above the effect-size ceiling reported for functional connectivity in the same data. The marker is not computed from raw functional connectivity but read out from a whole-brain generative model fitted to each individual’s dynamics, which gives access to interference structure that covariance-based features cannot represent. The regions driving the prediction, including precuneus, ventromedial prefrontal and anterior cingulate cortices, are among those previously implicated in internalising disorders, and the same signature tracks cognitive variation in healthy participants and is mechanistically linked to the efficiency of task-related computation. These findings establish a mechanistically interpretable and prospectively predictive marker of adolescent mental health and define a clear path towards external validation and clinical use.

## Introduction

Adolescence is a critical period of great promise and upheaval, characterised by significant behavioural and neurobiological changes (Sawyer *et al*., 2018), and increased vulnerability to mental health conditions (Paus *et al*., 2008; Solmi *et al*., 2022) including depression and anxiety (Kessler *et al*., 2005; Rapee *et al*., 2019; Solmi *et al*., 2022). The underlying reasons for this increased vulnerability remain poorly understood, but this is likely to change with recent large-scale neuroimaging initiatives such as the Human Connectome Project Boston Adolescent Neuroimaging of Depression and Anxiety (HCP BANDA) (Hubbard *et al*., 2020). Promising progress has been made using connectome-based predictive modelling with functional connectivity, with recent work in the BANDA cohort yielding a one-year prediction correlation of ρ = 0.22 (Morfini *et al*., 2025). This result is consistent with the broader effect-size ceiling for brain–behaviour associations reported across large neuroimaging consortia (Marek *et al*., 2022), and raises the question of whether a fundamentally different class of features, derived from fitted dynamical models of individual brain dynamics rather than from raw covariance, can move beyond that ceiling.

Early detection of problems with adolescent mental health is fundamental for enabling prompt and effective treatment (McGorry and Mei, 2018; Pizzagalli, 2022). Here, we were inspired by the historic use of sentinel animals such as canaries for obtaining early warning signals of toxic gases, primarily carbon monoxide in mines. Similarly, early-warning signals for mental health problems must be linked to deviations from healthy brain dynamics, which are fundamentally characterised by efficiency in speed and energy consumption (Deco *et al*., 2025c; Kringelbach *et al*., 2024). In fact, energy, information, and non-equilibrium are the three pillars of human brain dynamics responsible for our remarkable computational power while operating within a limited energy budget (Balasubramanian, 2021; Levy and Calvert, 2021). Thus, a framework linking these elements dynamically could serve as a sensitive sentinel for early detection of deviations in brain dynamics associated with mental health.

Promising new research has shown that the efficiency of the human brain arises from brain oscillations giving rise to long-range interactions through interference effects characterised by interference structure across distant regions (Deco *et al*., 2025b). The emergent states are produced by cluster synchronisation of systems of coupled oscillators using whole-brain modelling of empirical brain data (Deco and Kringelbach, 2025; Deco *et al*., 2017; Kringelbach and Deco, 2020; Pope *et al*., 2023).

Hence, we developed a novel framework to act as a ‘canary in the mind’ for early detection of mental health problems in adolescence. This framework extracts signatures of entanglement in brain dynamics based on interference. This is based on recent advances in physics where QL (quantum-like; contextual interference-based) effects can be found at the macroscopic level in systems of non-quantum (classical) coupled oscillators (Scholes, 2024, 2025; Scholes and Amati, 2025), including in brain dynamics where they are consistent with a mechanistic role for efficient processing (Deco *et al*., 2025b). Note this is not quantum physics in the microscopic sense but instead a probabilistic framework using the mathematics of quantum physics to capture classical interferences. Importantly, this signature of entanglement is computed not on the empirical timeseries directly but on the fitted whole-brain model that has already absorbed the empirical covariance and time-shifted covariance into its generative structure. This denoising step, together with the access it provides to interference effects that covariance cannot represent, is what allows the feature to carry information beyond what functional connectivity alone can deliver. The architectural framework that motivates measuring entanglement in brain dynamics, in which emotion, quantum-like binding inference and hierarchical orchestration are derived from a single thermodynamic constraint, is set out in a companion preprint (Kringelbach *et al*., 2026). The mathematical scaffolding showing that the structural and functional measures of entanglement used here are readings of one self-adjoint connectome operator is set out in a second companion preprint (Kringelbach and Deco, 2026).

Here, we define structural and functional measurements of entanglement. We use the structural measure of ‘spectral gaps’, which characterises algebraic connectivity and associated emergent states, acting as the structural skeleton underlying interference, i.e., intrinsically metastable cluster synchronisation configurations that give rise to the rich dynamical repertoire of the brain. This repertoire can be quantified using turbulence as a measure of metastable cluster synchronisation over time (Deco and Kringelbach, 2020; Deco *et al*., 2023; Deco *et al*., 2025a; Escrichs *et al*., 2024; Escrichs *et al*., 2022; Martinez-Molina *et al*., 2024). We define a turbulent vortex space, which contains the instantaneous levels of local synchronisation (Deco *et al*., 2025c). From this, a natural measure of entanglement in brain dynamics emerges as the Växjö Interference Connectivity (VIC) matrix, quantifying interference in brain dynamics as the violation of the law of total probability in the turbulent vortex space (Bulinski and Khrennikov, 2004; Khrennikov, 2021).

Using this entanglement of brain dynamics together with state-of-the-art connectome-based predictive modelling, we used the HCP BANDA individual’s baseline resting-state fMRI data to predict the one-year follow-up RCADS scores (r = 0.60). To better understand why this framework is so accurate for prediction in disease, we further investigated the underlying mechanisms of entanglement in relation to computational efficiency in health. In addition to providing support for potential causal mechanisms, we show that, in a large-scale HCP dataset, entanglement also significantly infers complex behavioural scores such as general intelligence (r = 0.79).

This paper is one of three companion preprints that together set out a mathematical unification of brain dynamics and its implications for the architecture of conscious experience. The single-operator unification (Kringelbach and Deco, 2026) demonstrates that connectome harmonics, the turbulence smoothing kernel and the complex harmonics form are three readings of one self-adjoint operator under the functional calculus, an identity tested with an LSD perturbation in which a single scalar coupling parameter predicts shifts in two mathematically independent functional domains. The Entangled Loop (Kringelbach *et al*., 2026) is the architectural theory this unification scaffolds, in which the primacy of emotion, the quantum-like character of binding inference and the hierarchical orchestration that coordinates them are derived from a single thermodynamic constraint. The present paper reports a first application of that framework, with external replication identified as the immediate next step.

## Results

We developed a new framework for measuring entanglement in brain dynamics to address the major challenges of predicting clinical outcomes from resting-state neuroimaging. In the following, we first describe the overall entanglement framework, then demonstrate why this framework is both sensitive and robust by investigating large-scale healthy cohorts. We then apply this framework to the HCP BANDA dataset, which contains resting-state neuroimaging at baseline and clinical RCADS (Revised Children’s Anxiety and Depression Scale) at both baseline and one-year follow-up. After showing how the framework predicts clinical outcomes, we demonstrate that it can also be used to infer behavioural scores in healthy participants, in particular the g-factor of intelligence. Finally, we provide a mechanistic explanation of why the framework is successful by linking it to the concept of computability.

### Overall framework for measuring entanglement in brain dynamics

Figure 1. shows a schematic overview of the framework, including its individual components, from preprocessing of neuroimaging and clinical data (**Figure 1A**), to the analysis pipeline used to extract the key measure of entanglement in brain dynamics for each individual (**Figure 1B**), and finally to the prediction of clinical outcomes (**Figure 1C**).

**Figure 1.**
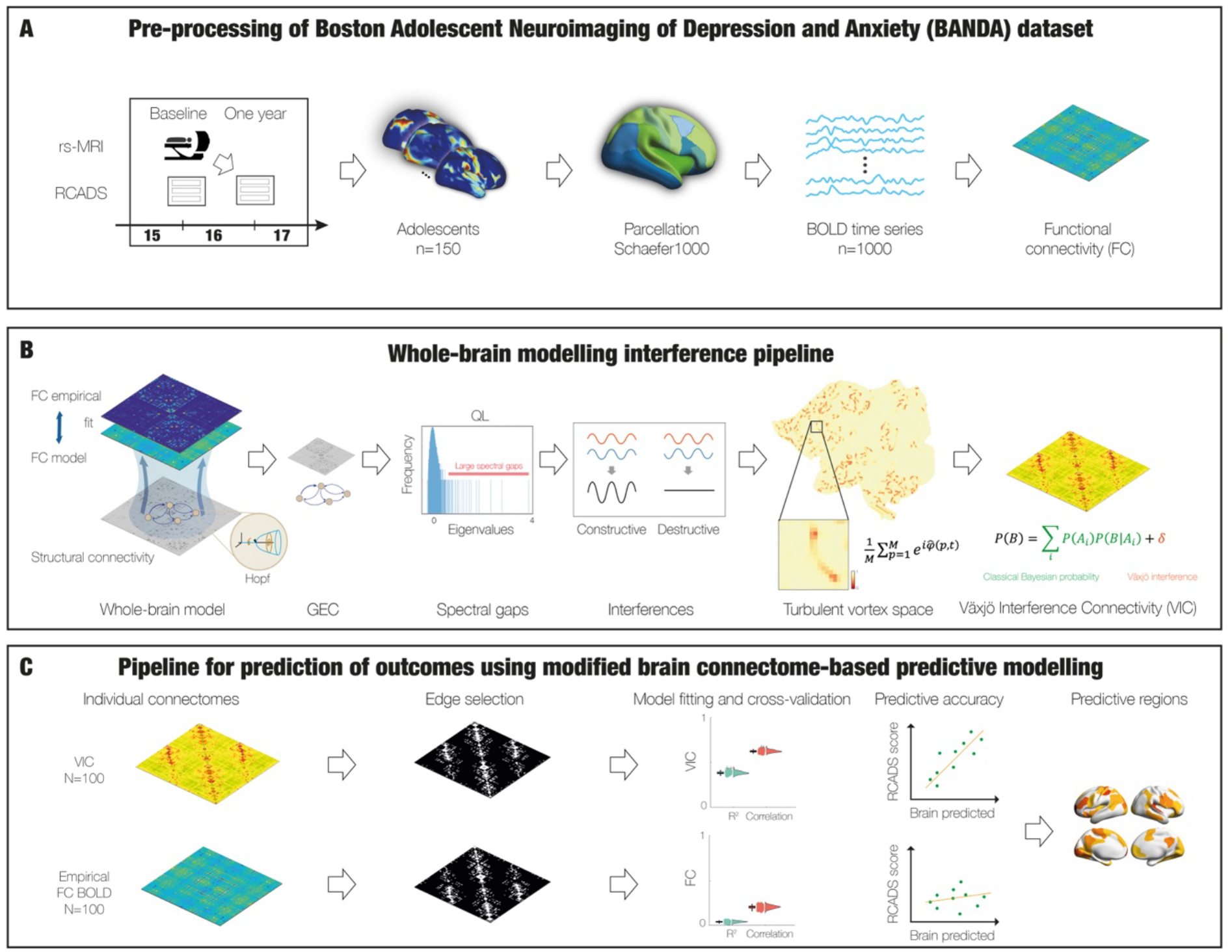
Framework for measuring entanglement in brain dynamics. **A)** The HCP BANDA (Boston Adolescent Neuroimaging of Depression and Anxiety) dataset contains resting-state neuroimaging at baseline and clinical RCADS (Revised Children’s Anxiety and Depression Scale) at both baseline and one-year follow-up. We created individual functional connectivity (FC) matrices from the extracted time series in the Schaefer1000 parcellation. **B)** We computed the individual Växjö Interference Connectivity (VIC) matrix using a whole-brain modelling pipeline for each individual. First, we created a Hopf whole-brain model of the Schaefer1000 data, resulting in a GEC (Generative Effective Connectivity) matrix fitting the empirical data (0.73 +/-0.05). Then we computed the spectral gaps of this GEC as a measure of QL computation. Following this, we computed the turbulent vortex space, i.e. instantaneous levels of local synchronisation in a coarser parcellation (Schaefer100) turbulent vortex space across time. This allowed us to measure the ‘entanglement of brain dynamics’ as the VIC, which is the interference given by the violation of the law of total probability in vortex space. **C)** Using state-of-the-art modified Connectome-based Predictive Modelling, we followed the canonical pipeline with both the VIC and FC matrices to predict the clinical one-year follow-up RCADS scores. The relevant brain networks predicting these scores can be rendered.

In terms of the clinical data, we used the HCP BANDA dataset with resting-state neuroimaging at baseline and clinical RCADS at both baseline and one-year follow-up (Hubbard *et al*., 2020; Siless *et al*., 2020). We extracted the time series using the Schaefer1000 atlas (Schaefer *et al*., 2018) and computed individual functional connectivity (FC) matrices for use in a modified connectome-based predictive modelling framework (mCPM) (Greene *et al*., 2018; Shen *et al*., 2017).

To define and extract the individual entanglement measure, we first used whole-brain modelling (Deco and Kringelbach, 2025) to identify the model best fitting the empirical data. This yields the Generative Effective Connectivity (GEC) matrix, an individually optimised version of the anatomical structural connectivity (SC), in which each pairwise coupling is assigned an optimal effective conductivity through an iterative procedure.

As specified in the *Methods*, we used the additional optimisation provided by our thermodynamic framework, incorporating both FC and time-shifted FC in fitting the whole-brain model (Kringelbach *et al*., 2024; Kringelbach *et al*., 2023). This breaks the time symmetry of FC and generates an asymmetric, hierarchical GEC, resulting in an excellent fit to the empirical BANDA data (0.73 ± 0.05).

This allows us to define a specific measure of entanglement based on both structural and functional properties. First, we extract the structural measure of ‘spectral gaps’, which characterises algebraic connectivity and associated emergent states. These form the structural backbone underlying intrinsically metastable cluster synchronisation configurations that collectively generate the rich dynamical repertoire of the brain.

For the functional measure, we use turbulence as a proxy for metastable cluster synchronisation over time. Briefly—see *Methods* for full details—this defines the turbulent vortex space, representing instantaneous levels of local synchronisation. This allows us to define a measure of entanglement in brain dynamics as VIC matrix. The VIC quantifies interference in brain dynamics as the violation of the law of total probability within the turbulent vortex space (Bulinski and Khrennikov, 2004; Khrennikov, 2021).

Finally, we use the mCPM framework to compare the predictive performance of VIC and FC matrices for the clinical one-year follow-up RCADS scores.

### Entanglement of brain dynamics in healthy participants

Using large-scale 3T neuroimaging data from the Human Connectome Project dataset (971 participants), we demonstrate that human brain dynamics exhibit both structural and functional entanglement linked to QL computation.

**Figure 2A** shows spectral gaps as a structural measure of QL computation across different connectomes. From right to left, the figure shows spectral gaps for: 1) the optimal GEC, 2) GEC without long-range connections, 3) the initial anatomical structural connectivity (SC; at the beginning of optimisation after 20 iterations for the individual GEC), and 4) a shuffled version of the SC (shuffled edge positions 100 times). The results clearly show that the GEC exhibits significantly larger spectral gaps than all other cases, demonstrating its suitability for QL computation and functional entanglement. The two panels of **Figure 2B** illustrate examples of small spectral gaps for the SC and large spectral gaps for the fully optimised GEC.

**Figure 2.**
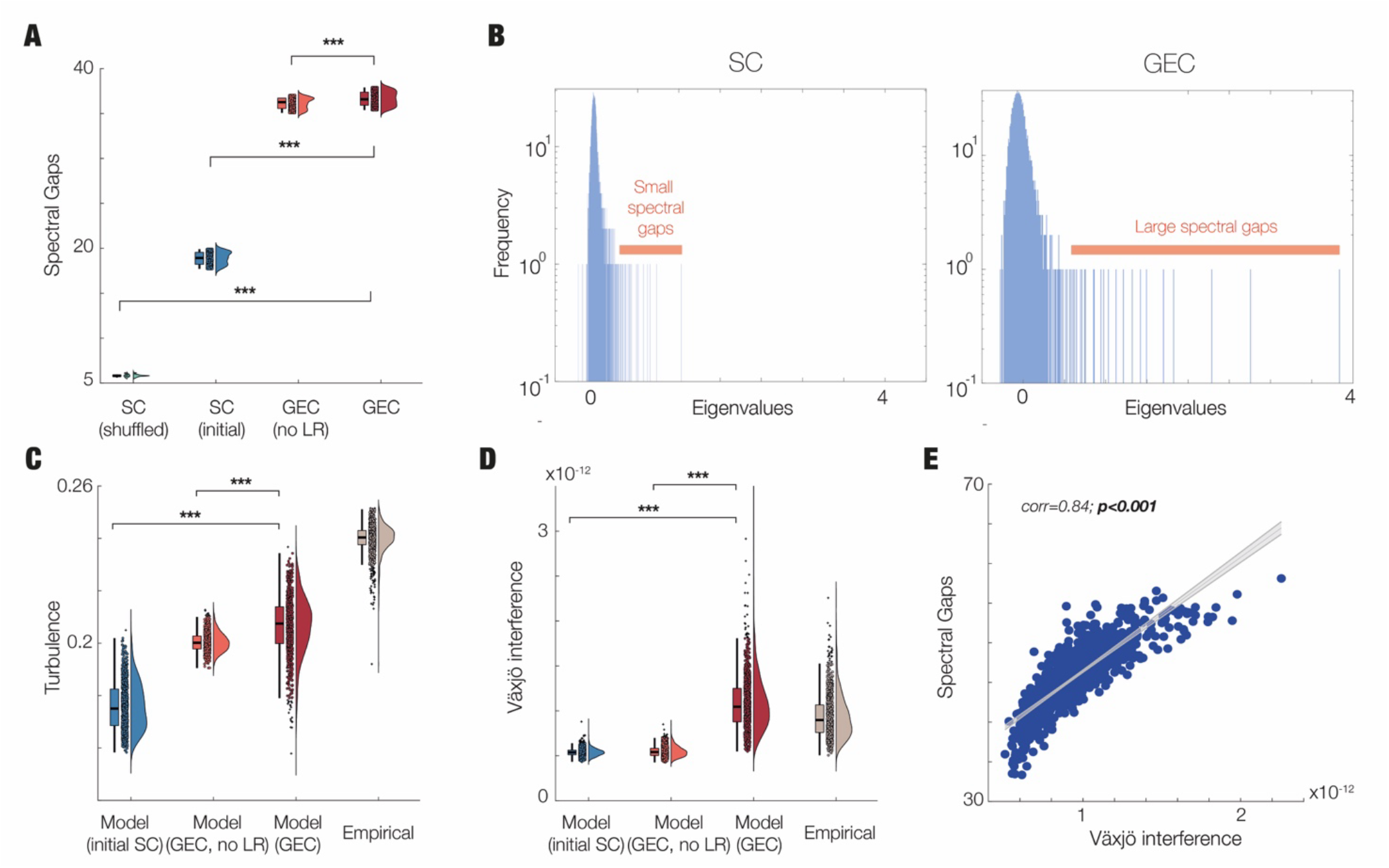
Robustness of entanglement of brain dynamics in healthy large scale human neuroimaging. We used two independent structural and functional measures of quantum-like (QL) computation in human brain dynamics obtained in the Human Connectome Project Dataset of 971 participants. **A)** First, we used spectral gaps as a measure of QL computation in structural connectomes. The figures show the spectral gaps for four different connectomes (from right to left): 1) the optimal Generative Effective Connectivity (GEC), corresponding to the best fitting of the empirical data, 2) GEC without long-range connections (no LR), 3) the initial anatomical structural connectivity (SC, at the beginning of optimisation for the individual GEC) and 4) a shuffled version of the SC. This shows that the GEC has significantly higher spectral gaps than any of the other cases, demonstrating QL computation in the human brain. **B)** We show examples of the spectral gaps for the SC and for the fully optimised GEC. **C)** In terms of functional measures of QL, we first show that turbulence reflects the underlying QL computation for the functional timeseries generated by the model, whether generated by SC, GEC without LR or optimal GEC. In all cases, the GEC model shows significantly higher turbulence than the other two and closer to the empirical data (on the right). **D)** Second, we show that Växjö interference (as a direct measure of QL computation) shows exactly the same pattern where the model GEC is close to the empirical data but significantly different from the other cases. **E)** We found that both spectral gap and Växjö interference significantly correlate across 971 HCP participants.

Turning to functional measures of entanglement, **Figure 2C** shows that turbulence reflects the underlying QL computation in the functional time series. We measured turbulence for empirical data and for time series generated by whole-brain models based on SC, GEC without long-range connections, and optimal GEC (as in **Figure 2A**). The results show that the GEC model exhibits significantly higher turbulence than the other connectomes and more closely matches the empirical data, confirming that large spectral gaps support metastable cluster synchronisation over time.

Importantly in **Figure 2D**, and as a direct functional measure of entanglement, we show that VIC – introduced as a measure of QL computation (Khrennikov, 2021) – exhibits the same pattern of results as turbulence. Specifically, the GEC model closely matches empirical data and differs significantly from the other connectomes. **Figure 2E** confirms this by showing that individual measures of spectral gap and VIC are significantly correlated across 971 HCP participants.

Overall, these results show that human brain dynamics can be described in terms of entanglement: the structure of the connectome exhibits large spectral gaps that support a rich repertoire of metastable cluster synchronisation configurations, captured functionally by VIC.

### Using entanglement in brain dynamics for improving prediction of clinical outcomes

Having established that healthy brain dynamics exhibit high levels of entanglement as measured by VIC, we hypothesised that this is compromised to varying degrees in neuropsychiatric conditions. To test this hypothesis, we applied the entanglement framework to the HCP BANDA dataset, as illustrated in **Figure 1**.

Given the relatively short resting-state time series in this dataset, we constructed optimised individual whole-brain models and used these to generate VIC matrices for each adolescent. **Figure 3A** shows the results of modified connectome-based predictive modelling (*see Methods*), yielding a significant correlation of r = 0.60 (p < 0.001) between empirical and predicted one-year follow-up RCADS scores.

**Figure 3.**
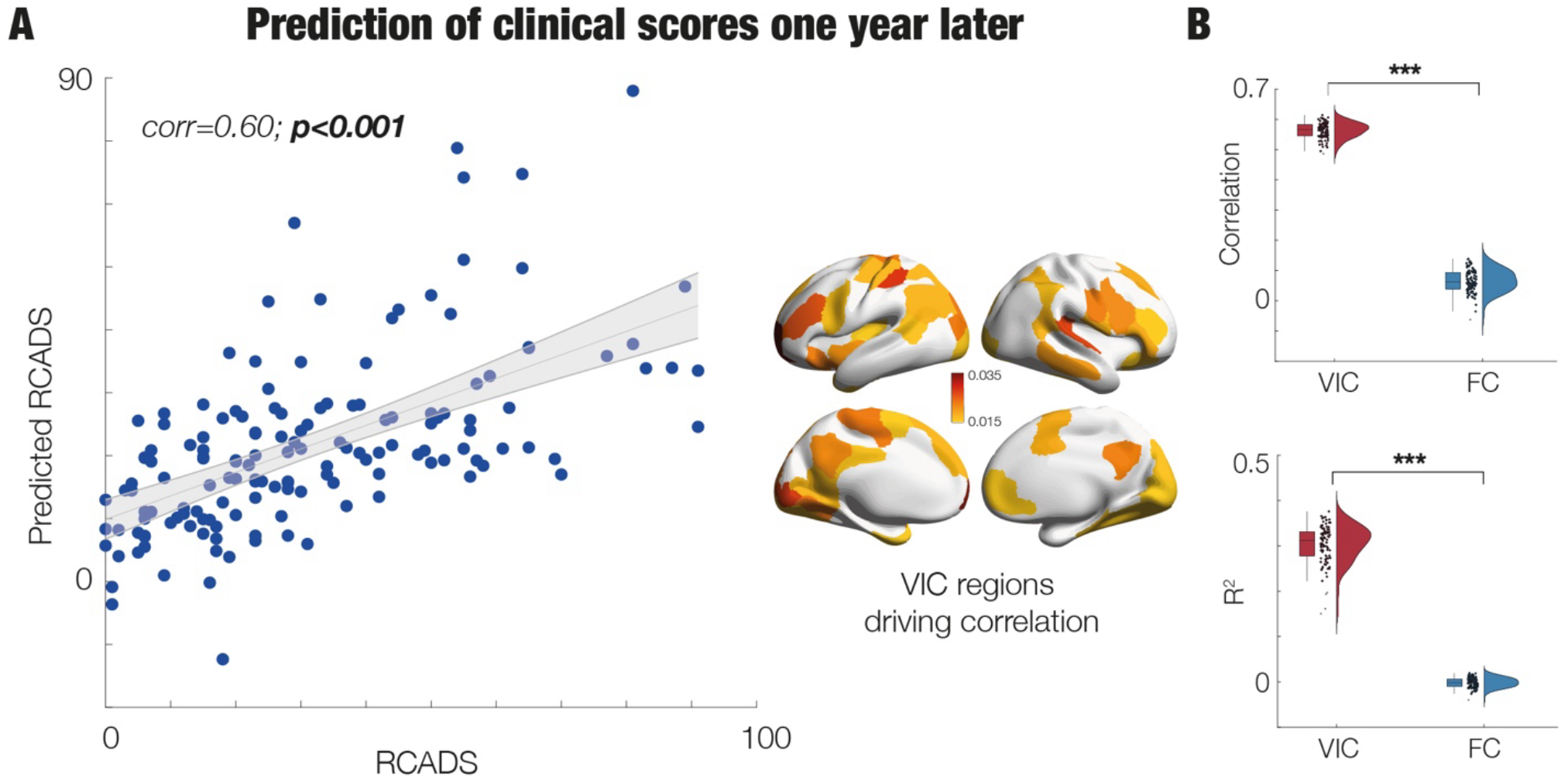
Entanglement of brain dynamics is an excellent predictor for clinical scores both at baseline and one-year follow-up. **A)** Using connectome-based predictive modelling with the individual Växjö Interference Connectivity (VIC) matrices yields significant prediction with r = 0.60 (p<0.001) between empirical and predicted one-year follow-up RCADS scores. The rendering of brains is showing the VIC of the top brain regions responsible for predicting the RCADS scores. **B)** The boxplots show the significant differences in predictability for VIC compared to functional connectivity (FC) with correlation on the top boxplot and R^2^ on the bottom boxplot.

Using VIC as input features significantly improved prediction compared to FC, as shown in both correlation (**Figure 3B**, top right boxplot) and R^2^ (bottom right boxplot). We also provide a rendering of VIC-based brain regions driving the predicted RCADS scores. These values are computed from the average centrality of mCPM-selected features across the respective participant groups.

### Entanglement infers intelligence and other behavioural scores in healthy participants

Given the strong predictive performance of entanglement-based measures, we applied VIC to a well-known challenging problem in neuroimaging: Inferring general intelligence (g-factor). For this analysis, we used 182 HCP participants scanned with high-quality 7T resting-state data.

**Figure 4A** shows that mCPM using VIC yields a significant correlation of r = 0.79 (p < 0.001) between inferred and empirical g-factor scores (left panel). In contrast, mCPM using FC produces a lower correlation (r = 0.53; right panel). The middle panel shows that VIC significantly outperforms FC in both correlation and R^2^.

**Figure 4.**
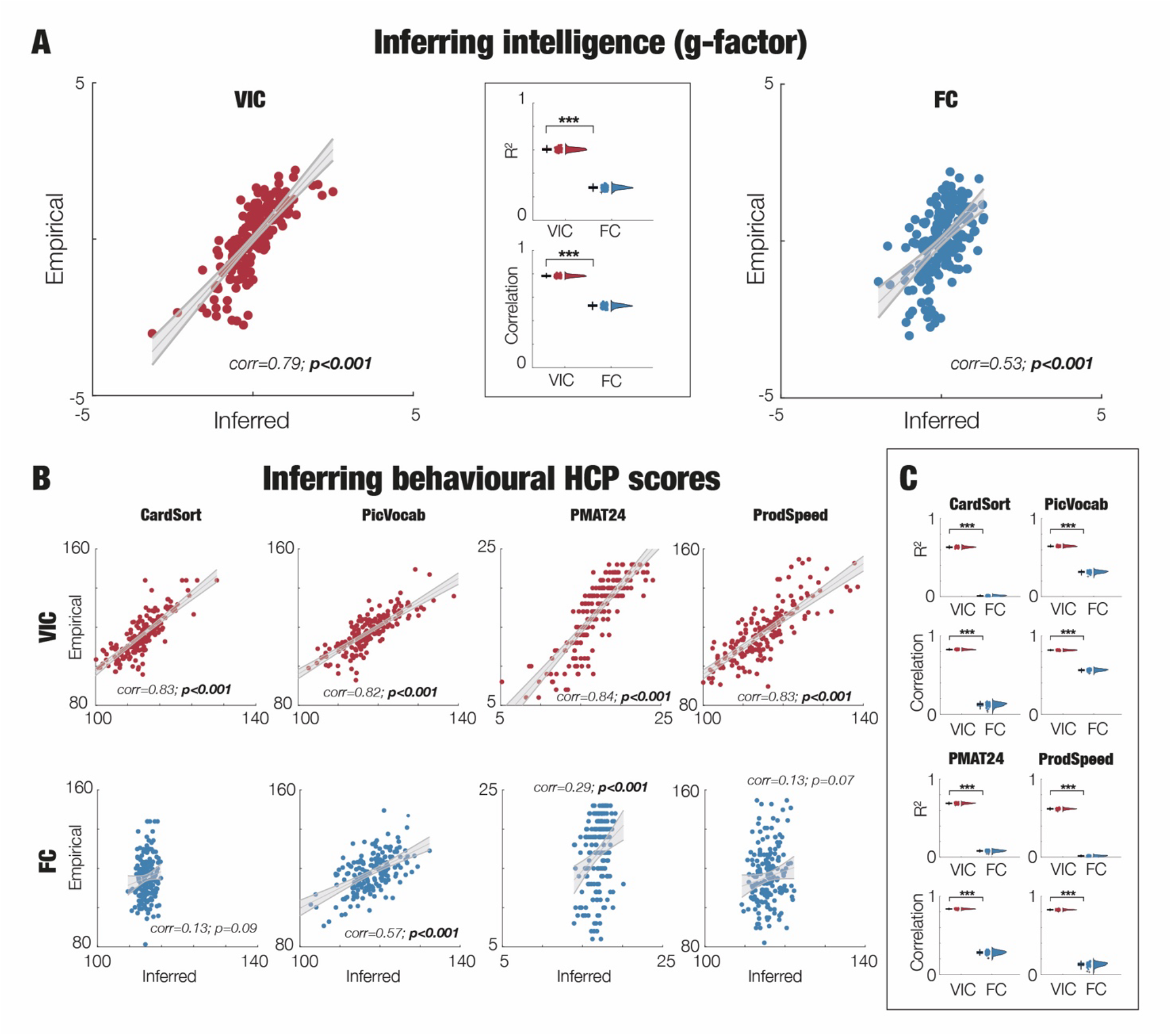
Entanglement of brain dynamics is also excellent for inferring intelligence and related behavioural scores in healthy participants. **A)** Applying a modified Connectome-based Predictive Modelling (mCPM) with VIC yields a significant correlation of 0.79 (p<0.001) between inferred and empirical g-factor scores across participants (left panel). While FC can also be used with mCPM to infer g-factor (right panel), the correlation is lower (0.53) and lower than using this with VIC where both correlation and R^2^ are significantly higher (see middle panel inset with the two boxplots). **B)** Similarly, the same is observed with the main scores in different cognitive domains (CardSort, PicVocab, PMAT24 and ProdSpeed), where all scores are significantly inferred with VIC, and where the correlation scores are always higher than for FC (CardSort: 0.83 vs 0.13; PicVocab: 0.82 vs 0.57, PMAT24:0.84 vs 0.29 and ProdSpeed:0.83 vs 0.13). **C)** This is even more clear when comparing the boxplots for the correlation and R^2^ in the inference (all boxplots are significantly higher for VIC vs FC).

We further evaluated predictive performance across multiple cognitive domains (CardSort, PicVocab, PMAT24, and ProdSpeed). **Figure 4B** shows that all four scores are significantly inferred using VIC, with consistently higher correlations than FC (CardSort: 0.83 vs 0.13; PicVocab: 0.82 vs 0.57; PMAT24: 0.84 vs 0.29; ProdSpeed: 0.83 vs 0.13).

**Figure 4C** further confirms that VIC consistently outperforms FC, with significantly higher correlation and R^2^ values across all behavioural inferences.

### Entanglement underlies optimal computability cost and energy

To understand why entanglement is highly predictive of behaviour from brain dynamics, we investigated its causal relationship with computability. As we have shown elsewhere, human brain computation depends on the dynamic contribution of neuromodulation, which allows apparently fixed anatomical connectivity to be flexibly reconfigured depending on task or state demands (Deco *et al*., 2026).

**Figure 5A** illustrates how we constructed whole-brain models for individual healthy participants and examined how neuromodulation, based on 19 neurotransmitter receptors and transporters, enables transitions from resting-state to different task demands. We define *computability* as the ability of the brain to flexibly transition between resting-state and all seven tasks.

**Figure 5.**
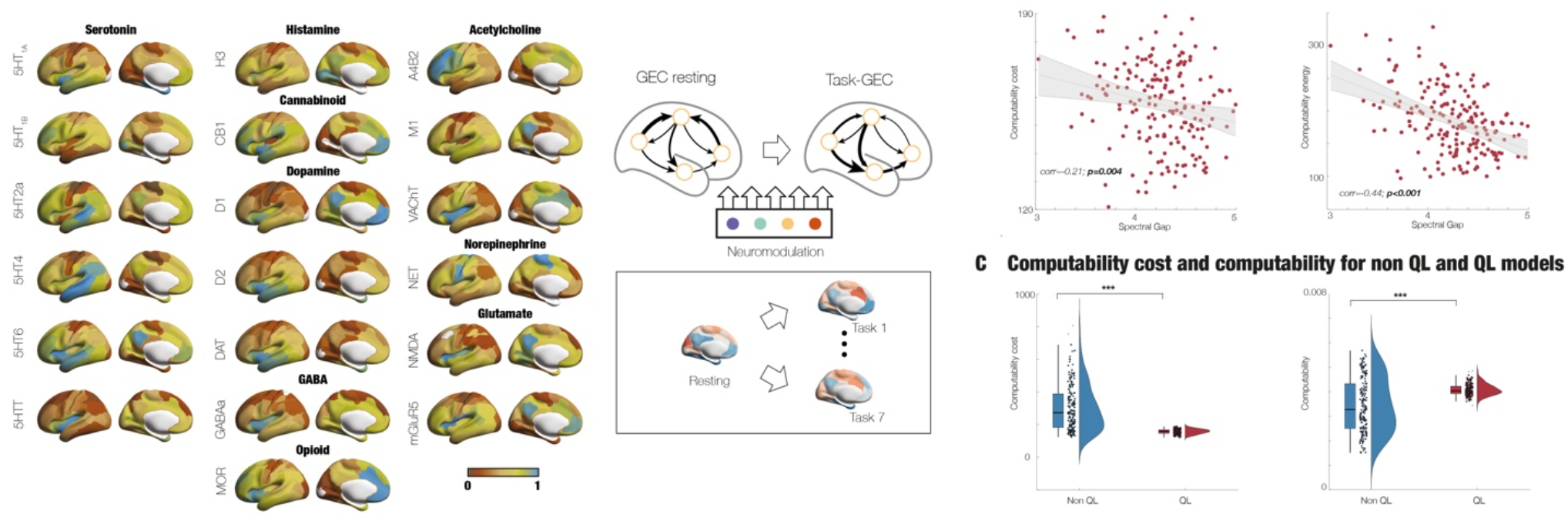
Entanglement of brain dynamics has lower computability cost and computability energy compared to when this is diminished in an otherwise equivalent whole-brain model. **A)** Computation in the human brain depends on the contribution of neuromodulation to dynamically change its function depending on task or state demands. Here, we used 19 different neurotransmitter receptors and transporters across nine different neurotransmitter systems mapped with PET tracer images from more than 1,200 healthy participants collated and averaged to produce mean receptor distribution maps (Hansen et al., 2022). We defined ‘computability cost’ as the weighted contribution of these 19 maps changing the individual optimal GEC resting to best fitting the task (task-GEC). We do this for all seven tasks in the HCP dataset. As such, ‘computability’ is defined as the ability of the brain to flexibly change between resting and all seven tasks. Equally, the ‘computability energy’ is the thermodynamic energy cost of switching from resting to a task state (see Methods). **B)** The scatterplots show the significant correlations between the GEC spectral gaps and the computability cost (left panel) and computability energy (right panel). This demonstrates that quantum-like (QL) computation facilitates computation. **C)** Similarly, when creating models that are either non-QL or QL, the computability costs (left panel) are significantly lower and computability (right panel) is significantly higher for QL. Again, this demonstrates that QL is fundamental for efficient and flexible computation.

We define *computability cost* as the weighted contribution of these 19 neurotransmitter maps required to transform the individual optimal resting-state GEC into the task-specific GEC (task-GEC), across all seven HCP tasks. Similarly, *computability energy* is defined as the thermodynamic energy cost associated with switching from resting-state to a task state (Deco *et al*., 2025d).

The results are shown in **Figure 5B**. The scatter plots reveal significant correlations between GEC spectral gaps and both computability cost (left panel) and computability energy (right panel). These findings provide direct evidence that QL computation facilitates efficient computation in terms of both energy and flexibility.

As shown in **Figure 5C**, we further investigated computability using whole-brain models constructed to exhibit either non-QL or QL dynamics. The box plots show that QL models exhibit significantly lower computability costs (left panel) and significantly higher computability (right panel) compared to non-QL models.

Together, these results provide strong evidence that entanglement and QL dynamics are directly linked to efficient and flexible computation in the healthy human brain.

## Discussion

Here we propose a ‘sentinel’ framework to address a major challenge in neuroimaging, namely predicting behaviour (including long-term emotional symptoms) from brain signals. We demonstrate how our ‘canary in the mind’ framework, based on a measure of entanglement in brain dynamics, can accurately predict clinical behavioural scores in adolescents. This approach has the potential to improve outcomes by enabling targeted early intervention during critical developmental periods. Specifically, we show that baseline resting-state fMRI data can significantly predict the clinical one-year follow-up RCADS scores (r = 0.60, *p<0*.*001*) in the HCP BANDA cohort. Interestingly, as shown in **Figure 3**, the top brain regions driving this prediction are found mainly in regions such as the precuneus, ventromedial prefrontal and anterior cingulate cortices, which have previously been identified as orchestrating drivers of the transition to mental health problems (Deco *et al*., 2024; Kringelbach, 2005) and have shown robust and reproducible FC abnormalities in adults with major depressive disorder (Kaiser *et al*., 2015). These regions overlap with those the architectural framework identifies as the orchestrating ensemble through which valenced content is broadcast, in particular the precuneus and the cingulate cortex (Deco *et al*., 2021; Kringelbach *et al*., 2026).

The magnitude of this prediction warrants framing against the broader literature. Marek and colleagues showed that brain-wide association studies relying on functional connectivity features typically yield correlations in the range of 0.06 to 0.16 even in samples of thousands, with multivariate methods achieving higher but still modest effects (Marek *et al*., 2022). The result reported here sits well above that ceiling, but it is computed on features derived from a fitted dynamical model rather than from raw covariance, and so it is not situated in the same reference class. This distinction between raw measurements and model-derived quantities is well established in other fields. In genetics, polygenic risk scores aggregated through fitted statistical models predict phenotypes far more accurately than any individual locus, even though the loci themselves carry the ultimate signal (Khera *et al*., 2018). In climate science, ensemble projections from coupled physical models predict regional outcomes more reliably than direct extrapolation of station data (Hausfather *et al*., 2020; Tebaldi and Knutti, 2007). In both cases the model is not an additional layer on top of the data but a constraint that uses prior knowledge of the system’s dynamics to extract signal that the raw measurements carry but, critically, do not expose. The whole-brain Hopf model plays the same role here: By absorbing the empirical covariance and time-shifted covariance into a generative structure, it makes accessible the interference patterns that an unfitted covariance matrix cannot represent. The model-based interference structure captured by VIC is, on this view, the natural feature class for clinical prediction from brain dynamics.

The predictive performance of this framework arises from its ability to capture interference in brain dynamics, providing a sensitive measure of information flow. Using large-scale neuroimaging data, we show that the framework robustly captures both structural and functional measures of entanglement. Structurally, spectral gaps in the GEC are significantly larger than in the anatomical SC, suggesting that this feature enables the brain to exploit interference effects to generate a rich repertoire of cluster synchronisation states required for efficient computation.

At the functional level, this rich repertoire is reflected in the significantly higher turbulence observed in the GEC-based whole-brain model compared to the SC-based model. Consistent with this, we show that Växjö interference – a direct measure derived from QL systems – is also higher in the GEC model. Importantly, these interference effects arise in a classical system of coupled Hopf oscillators, supporting recent evidence that QL computation may underlie the speed and energy efficiency of healthy brain dynamics (Deco *et al*., 2025b).

Previous work has suggested that neuropsychiatric disorders may arise from disruptions in regions that coordinate brain dynamics at higher levels of the functional hierarchy (Deco *et al*., 2024). In this context, entanglement promotes interference across regions and thereby shapes hierarchical brain organisation (Kringelbach *et al*., 2026). Deviations from an optimal entangled state—and thus from efficient QL computation—may therefore lead to dysfunctional brain dynamics, which over time can contribute to neuropsychiatric disorders. This interpretation aligns with the metaphor of a ‘canary in the mind’: early deviations from healthy entanglement can signal emerging dysfunction before it becomes clinically severe.

To further support the mechanistic interpretation of our results, we introduced the concept of computability cost, defined as the cost required for transitioning from resting-state to task states. Using seven HCP tasks spanning cognitive and emotional domains, we identified optimal configurations of 19 neurotransmitter receptor maps across participants. We found significant correlations between GEC spectral gaps and both computability cost and computability energy. These findings suggest that QL computation in the healthy brain plays a key role in enabling efficient and flexible computation. The mathematical reason why the spectral gap controls computability simultaneously with turbulence and quantum-like interference, namely that all three are readings of one self-adjoint operator under the functional calculus, is established in a companion preprint (Kringelbach and Deco, 2026).

Consistent with this, whole-brain models exhibiting QL dynamics show significantly lower computability cost and higher computability compared to non-QL models. This provides mechanistic evidence linking entanglement, QL dynamics, and computational efficiency. The current discovery that a breakdown across these constructs predicts depressive/anxiety symptoms one year later is a novel and clinically important finding. Further demonstrating the generality of this framework, we show that entanglement-based measures also significantly infer intelligence and related behavioural scores in healthy participants.

Future clinical research along the lines outlined here would benefit from addressing two important caveats. First, the clinical prediction is based on internal cross-validation in a single cohort of 150 adolescents, and external replication on an independent longitudinal dataset will be needed to establish generalisation across sites and acquisition protocols. The use of different symptom instruments across available paediatric cohorts complicates direct cross-cohort prediction, but partial replications testing whether the predictive network generalises across diagnostic subgroups within BANDA are a natural next step. Second, while the mechanistic account linking entanglement to computability provides a principled basis for the predictive performance reported here, that account itself rests on whole-brain modelling assumptions that, although well-supported in healthy cohorts, will benefit from further validation in clinical populations.

Overall, these findings emphasise the importance of oscillatory brain dynamics and, in particular, the role of interference as an intrinsic feature of coupled oscillatory systems. Interference appears to enrich the repertoire of available brain states, facilitating flexible switching required for task performance. When this flexibility is compromised, it may contribute to the emergence of neuropsychiatric disorders.

The present framework therefore offers both theoretical insight and practical potential. As a ‘canary in the mind’, entanglement-based measures may provide a sensitive tool for early detection of deviations in brain dynamics, opening the possibility for earlier and more effective intervention.

## Data Availability

All neuroimaging data used in this study are publicly available through standard data-sharing procedures. The HCP BANDA cohort, used for the primary clinical prediction analyses, is available through the NIMH Data Archive (https://nda.nih.gov/) as the BANDA 1.1 Data Release (Hubbard et al., 2024). The Human Connectome Project Young Adult data used for the robustness analyses (1200 participants at 3T and 182 at 7T) are available through ConnectomeDB (https://db.humanconnectome.org/). Both datasets require registration and a signed Data Use Agreement before download. The Schaefer1000 cortical parcellation used to extract regional time series is available through the CBIG repository (https://github.com/ThomasYeoLab/CBIG). Code implementing the whole-brain modelling pipeline, the Vaxjo Interference Connectivity computation and the modified Connectome-based Predictive Modelling analysis is publicly available at https://github.com/decolab/canary.

## Acknowledgements

G.D. is supported by Grant PID2022-136216NB-I00 funded by MICIU/AEI/10.13039/ 501100011033 and by “ERDF A way of making Europe”, ERDF, EU, Project NEurological MEchanismS of Injury, and Sleep-like cellular dynamics (NEMESIS) (ref. 101071900) funded by the EU ERC Synergy Horizon Europe, and AGAUR research support grant (ref. 2021 SGR 00917) funded by the Department of Research and Universities of the Generalitat of Catalunya. Y.S.P. is supported by the project NEurological MEchanismS of Injury, and Sleep-like cellular dynamics (NEMESIS) (ref. 101071900) funded by the EU ERC Synergy Horizon Europe. D.A.P. was partially supported by National Institute of Mental Health P50 MH119467 and R37 MH068376. M.L.K. is supported by the Centre for Eudaimonia and Human Flourishing (funded by the Pettit and Carlsberg Foundations) and Center for Music in the Brain (funded by the Danish National Research Foundation, DNRF117). The funders had no role in study design, data collection and analysis, decision to publish or preparation of the manuscript.

## Methods

### Healthy Human Connectome Project (HCP) 7T dataset

#### Ethics

The Washington University–University of Minnesota (WU-Minn HCP) Consortium obtained full, written informed consent from all participants, and research procedures and ethical guidelines were followed in accordance with Washington University institutional review board approval (Mapping the Human Connectome: Structure, Function, and Heritability; IRB # 201204036).

#### Participants

All data were extracted from the Human Connectome Project (HCP), including resting-state and task data from 971 participants (3T), and resting-state data from 182 participants (7T).

#### Experimental Paradigm: Resting and Task

All participants performed a resting-state and the standard HCP task battery described in detail on the HCP website (Barch *et al*., 2013). This consists of seven tasks: Working memory (WM), motor, gambling, language, social, emotional, relational. According to HCP, the tasks were designed to cover a broad range of human abilities in several major domains sampling the diversity of the brain: 1) visual, motion, somatosensory, and motor systems, 2) category-specific representations; 3) working memory, decision-making and cognitive control systems; 4) language processing; 5) relational processing; 6) emotion processing; and 7) social cognition. All HCP participants performed all tasks in two separate sessions (first session: gambling, WM and motor; second session: language, emotion processing, relational processing and social cognition).

#### Measuring the g-factor

The *g*-factor was computed using a Matlab adaptation of the procedure described by Dubois and colleagues (Dubois *et al*., 2018) to perform factor analysis of the scores on ten relevant cognitive scores from the behavioural psychometric battery used to assess each HCP individual. This procedure derives the *g*-factor measure of intelligence, which is a standard used in the field of intelligence research.

#### Neuroimaging Data

The HCP website (http://www.humanconnectome.org/) provides the full details of participants, the acquisition protocol and preprocessing of the data. Below we have briefly summarised this information.

#### 3T Structural Data

The HCP structural data were acquired using a customized 3 Tesla Siemens Connectome Skyra scanner with a standard Siemens 32-channel RF-receive head coil. For each participant, at least one 3D T1w MPRAGE image and one 3D T2w SPACE image were collected at 0.7 mm isotropic resolution.

#### 3T Diffusion MRI

In order to reconstruct a high-quality structural connectivity (SC) matrix, we obtained multi-shell diffusion-weighted imaging data from 32 participants from the HCP database (scanned for approximately 89 minutes). The acquisition parameters are described in detail on the HCP website (Setsompop *et al*., 2013). This is used for constructing the whole-brain model as the first estimate of the Generative Effective Connectivity (GEC), which is then iteratively improved to fit the functional data (see below).

#### 3T Functional Data

Briefly, the HCP 3T fMRI data was acquired using a customized 3 Tesla Siemens Connectome Skyra scanner with a standard Siemens 32-channel RF-receive head coil, with the following parameters: 2.0 mm isotropic voxels, TR = 720 ms, TE = 33.1 ms, flip angle = 52 degrees, FOV = 208 × 180 mm, 72 slices, multi-band factor = 8. For full details of the task scans, see http://protocols.humanconnectome.org/HCP/3T/imaging-protocols.html.

#### 7T Functional Data

For each participant, HCP fMRI data were acquired using a 7 Tesla Siemens Magnetom scanner with a Nova32 32-channel RF-receive head coil, using the following parameters: 1.6 mm isotropic voxels, TR = 1000 ms, TE = 22.2 ms, flip angle = 45 degrees, matrix = 130 x 130, FOV = 208 × 208 mm, 85 slices, multi-band factor = 5, image acceleration factor (iPAT) = 2, partial Fourier sampling = 7/8, echo spacing = 0.64 ms, bandwidth = 1924 Hz/Px. The direction of phase encoding alternated between posterior-to-anterior and anterior-to-posterior across runs. For full details, see http://protocols.humanconnectome.org/HCP/7T/.

#### Neuroimaging Preprocessing for fMRI HCP

The preprocessing of the HCP resting-state and task datasets is described in detail on the HCP website. Using an existing pipeline (Deco *et al*., 2022), the data were preprocessed using the HCP pipeline which relied on standardised methods using FSL (FMRIB Software Library), FreeSurfer, and the Connectome Workbench software (Glasser *et al*., 2013; Smith *et al*., 2013). This standard preprocessing included correction for spatial and gradient distortions and head motion, intensity normalization and bias field removal, registration to the T1 weighted structural image, transformation to the 2mm Montreal Neurological Institute (MNI) space, and using the FIX artefact removal procedure (Navarro Schroder *et al*., 2015; Smith *et al*., 2013). The head motion parameters were regressed out and structured artefacts were removed by ICA+FIX processing (Independent Component Analysis followed by FMRIB’s ICA-based X-noiseifier (Griffanti *et al*., 2014; Salimi-Khorshidi *et al*., 2014)). Preprocessed timeseries of all grayordinates are in HCP CIFTI grayordinates standard space and available in the surface-based CIFTI file for each participant for resting-state and each of the seven tasks.

We used a custom-made Matlab script using the ft_read_cifti function (Fieldtrip toolbox (Oostenveld *et al*., 2011)) to extract the average timeseries of all the grayordinates in each region of the Schaefer1000 atlas, which are defined in the HCP CIFTI grayordinates standard space. The BOLD timeseries were filtered using a second-order Butterworth filter in the range of 0.008-0.08Hz.

### HCP BANDA (Boston Adolescent Neuroimaging of Depression and Anxiety)

#### Ethics

Participating parents provided informed consent and adolescents assented to study procedures and data sharing. Study procedures and data sharing were approved by the Massachusetts General Brigham Institutional Review Board (Protocol #P002534). Parents and adolescents were compensated for their time.

#### Participants and Dataset

The data were collected from October 2016 to November 2021 and contains depressed-anxious and healthy adolescents (N = 215) ages 14–17 years. Data were obtained from the BANDA 1.1 Data Release (Hubbard *et al*., 2024). We only included the 150 participants with resting-state scans and with available symptom severity reported at the baseline and one-year follow-up, who had low-motion rs-fMRI data at baseline (FD<0.32).

#### MRI Acquisition

Briefly, the MRI data were collected on a Siemens 3T Prisma MRI with a 64-channel head coil. One high resolution anatomical image was acquired with a T1-weighted MPRAGE sequence (0.8 mm isotropic voxels, field of view = 256 x 240 x 167 mm, TR = 2,400 ms, TE = 2.18 ms). rs-fMRI were acquired using simultaneous multi-slice (2.0 mm isotropic voxels, 72 slices, multiband acceleration factor = 8, TR = 800 ms, TE = 37 ms, flip angle = 52^°^), each consisting of 420 volumes lasting 5 min and 46 s. Two sets of two eyes-open rs-fMRI (four total runs lasting overall ∼23) with opposite phase encoding (PE) direction (Anterior-Posterior [AP] and Posterior-Anterior [PA]) were acquired and were alternated with the acquisition of a Spin-Echo fieldmap with opposite PE direction (AP-PA).

#### fMRI Preprocessing

Functional MRI data were preprocessed using the standardised HCP minimal preprocessing pipelines (see above). Briefly, these procedures included gradient distortion correction, motion correction, EPI distortion correction, and alignment of functional images to the structural T1-weighted image. Data were subsequently normalised to a standard template and resampled to a common space. Additional processing steps typically included spatial smoothing, temporal filtering, and removal of nuisance signals such as motion parameters and physiological noise.

#### Behavioural Measures

Emotional and behavioural symptoms were assessed using the Revised Children’s Anxiety and Depression Scale (RCADS), a standardised questionnaire designed to measure symptoms of anxiety and depression in children and adolescents. The RCADS includes multiple subscales corresponding to major anxiety disorders and major depressive disorder. Items are rated on a Likert scale reflecting symptom frequency, and scores can be summarised as raw scores or standardised T-scores. Higher scores indicate greater levels of anxiety and depressive symptoms.

### Neuromodulation Data

#### Receptor maps from PET

Receptor densities were assessed using PET tracer studies for 19 different receptors and transporters across nine neurotransmitter systems from more than 1,200 healthy participants, based on data (available at GitHub) provided by Hansen and colleagues (Hansen *et al*., 2022). These neurotransmitter systems include dopamine (D1, D2, DAT, noradrenaline (NET), serotonin (5HT1A, 5HT1B, 5HT2A, 5HT4, 5HT6, 5HTT), acetylcholine (A4B2, M1, VAChT), glutamate (mGLUR5, NMDA), GABA (GABA-A), histamine (H3), cannabinoid (CB1), and opioid (MOR). The volumetric PET images were aligned to the MNI-ICBM 152 nonlinear 2009 (version c, asymmetric) template, averaged across participants within each study, and then parcellated. For receptors/transporters with multiple mean images from the same tracer (5HT1B, D2, VAChT), a weighted average was applied (Hansen *et al*., 2022).

### Whole-brain Dynamics

Mathematically, the whole-brain model uses dynamics as defined by

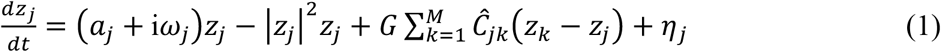

where for the oscillator in region *j*, the complex variable *z*_*j*_ denotes the state (*z*_*j*_ = *x* _*j*_ + i*y*_*j*_), *η*_*j*_ is additive uncorrelated Gaussian noise with variance *σ*^2^ (for all *j*), *ω*_*j*_ is the intrinsic node frequency, and *a*_*j*_ is the node’s bifurcation parameter. Within this model, the intrinsic frequency *ω*_*j*_ of each node is in the 0.008–0.08Hz band. The intrinsic frequencies were estimated from the empirical data, as given by the averaged peak frequency of the narrowband blood-oxygen-level-dependent (BOLD) signals of each brain region. For *a*_*j*_ > 0, the local dynamics settle into a stable limit cycle, producing self-sustained oscillations with frequency *ω*_*j*_/(2π). For *a*_*j*_ < 0, the local dynamics present a stable spiral point, producing damped or noisy oscillations in the absence or presence of noise, respectively. The fMRI signals were modelled by the real part of the state variables, i.e., *x* _*j*_ = Real(*z*_*j*_). The fitting of the whole-brain model is accomplished by finding the optimal fit of the global coupling parameter (global conductivity), *G*, by minimising the elementwise quadratic error between the functional connectivity matrices (Pearson correlations between the fMRI timeseries in each brain region) of the model (***FC***^*model*^) and empirical (***FC***^*emp*^) data.

This large-scale optimisation is made possible by the linearisation of the SL oscillator (Ponce-Alvarez and Deco, 2024). Proximity of the SL oscillator to criticality has been shown to provide the best working point for fitting whole-brain neuroimaging dynamics. This happens at the brink of the bifurcation, i.e. with *a*_*j*_ slightly negative but very near to zero (usually *a*_*j*_ = −0.02) (Deco *et al*., 2017). Briefly, we can estimate the functional correlations of the whole-brain network using a linear noise approximation (LNA). Hence, the dynamical system of *N* nodes (Equation 1) can be re-written in vector form as:

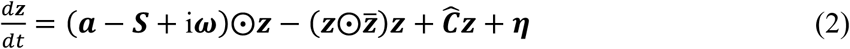

where ***z*** = [*z*_1_, …, *z*_*M*_]^*T*^, ***a*** = [*a*_1_, …, *a*_*M*_]^*T*^, ***ω*** = [*ω*_1_, …, *ω*_*M*_]^*T*^, ***η*** = [*η*_1_, …, *η*_*M*_]^*T*^ and ***S*** = [*S*_1_, …, *S*_*M*_]^*T*^ is a vector containing the strength of each node, i.e. 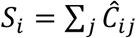 . The superscript []^*T*^represents the transpose, 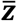 is the complex conjugate of ***z*** and ⨀ is the Hadamard element-wise product. As such, the equation describes the linear fluctuations around the fixed point ***z*** = 0, which is the solution of 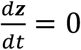 . Discarding the higher-order terms 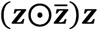 and separating the real and imaginary parts of the state variables, the evolution of the linear fluctuations follows a Langevin stochastic linear equation:

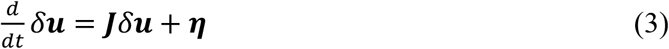

where the 2*M*-dimensional vector *δ****u*** = [*δ****x***, *δ****y***]^***T***^ = [*δx*_1_, …, *δx*_*M*_, *δy*_1_, …, *δy*_*M*_]^*T*^ contains the fluctuations of real and imaginary state variables. The 2*M* × 2*M* matrix ***J*** is the Jacobian of the system evaluated at the fixed point, which can be written as a block matrix

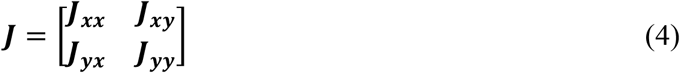

where ***J***_***xx***_, ***J***_***xy***_, ***J***_***yx***_, ***J***_***yy***_ are *M* × *M* matrices given as ***J***_***xx***_ = ***J***_***yy***_ = diag(***a*** − ***S***) + ***C*** and ***J***_***xy***_ = −***J***_***yx***_ = diag(***ω***), where diag(***v***) is the diagonal matrix whose diagonal is the vector ***v***. Please note that this linearisation is only valid if ***z*** = 0 is a stable solution of the system, that is if all eigenvalues of ***J*** have negative real parts. To compute the covariance matrix ***K*** = ⟨*δ****u****δ****u***^***T***^⟩, one can begin by writing Equation 3 as *dδ****u*** = ***J****δ****u****dt* + *d****W***, where *d****W*** is an 2*M*-dimensional Wiener process with covariance ⟨*d****W****d****W***^***T***^⟩ = ***Q****dt*, where ***Q*** is the noise covariance matrix, which is diagonal if the noise is uncorrelated. Using Itô’s stochastic calculus, we get *d*(*δ****u****δ****u***^***T***^) = *d*(*δ****u***)*δ****u***^***T***^ + *δ****u****d*(*δ****u***^***T***^) + *d*(*δ****u***)*d*(*δ****u***^***T***^). Noting that ⟨*δ****u****d****W***^***T***^⟩ = 0, taking the expectations and keeping terms to first order in the differential *dt*, we obtain:

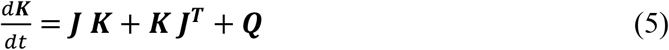

Hence, the stationary covariances (for which 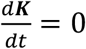) can be obtained by solving the following analytic equation:

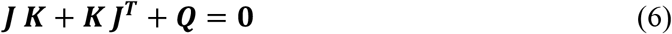

Equation 6 is a Lyapunov equation that can be solved using the eigen-decomposition of the Jacobian matrix (Deco *et al*., 2014). We obtained the simulated functional connectivity matrix ***FC***^*model*^ by reducing (through averaging across the *n* in each brain region) the functional connectivity obtained from the first *M* rows and columns of the covariance ***K***, which corresponds to the real part of the dynamics, and thus the BOLD fMRI signal.

#### Optimizing the Generative Effective Connectivity (GEC)

In order to create the generative effective connectivity (GEC), we used a pseudo-gradient procedure to optimise the initial coupling connectivity matrix ***C***, derived from the anatomical structural connectivity which was then iteratively used to create the GEC. The GEC is generative, and it is computed by using the whole-brain model to adapt the strength of the existing anatomical connectivity (i.e. the effective conductivity values of each fibre) to explain the functional data. For defining the GEC of a whole-brain model, we first fit the model to the empirical neuroimaging data for each participant. For this, we used a noise term *η*_*j*_ with variance *σ*^2^ = 0.02 homogeneous across all regions in a given parcellation

Specifically, we iteratively compared the output of the model with the empirical measures of the functional correlation matrix (***FC***^*empirical*^), i.e., the normalised covariance matrix of the functional neuroimaging data. Furthermore, we also compared the output of the model with the normalised *τ* time-shifted covariances (***FS***^*empirical*^(*τ*)). These normalised time-shifted covariance matrices are generated by taking the shifted covariance matrix ***KS***^*empirical*^(*τ*) and dividing each pair (*i, j*) by 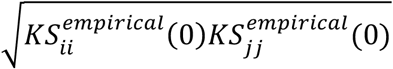.

Note that these normalised time-shifted covariances break the symmetry of the couplings and thus improve the level of fitting (Gilson *et al*., 2017). Using a heuristic pseudo-gradient algorithm, we proceeded to update the ***C*** until the fit is fully optimised. More specifically, the updating uses the following form:

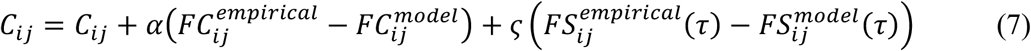

where 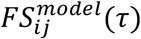 is defined similar to 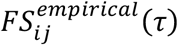. In other words, the updating is given by the first *N* rows and columns of the simulated *τ* time-shifted covariances ***KS***^*model*^(*τ*) normalised by dividing each pair (*i, j*) by 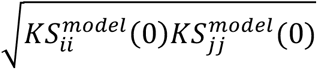, where ***KS***^*model*^(*τ*) is the shifted generative covariance matrix computed as follows:

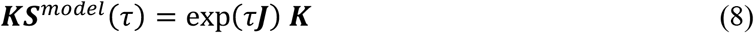

Please note that ***KS***^*model*^(0) = ***K***. The model was then run repeatedly with the updated ***C*** until the fit converges towards a stable value. As stated above, we initialised ***C*** using the average anatomical connectivity and only update known existing connections from this matrix (in either hemisphere). There is one exception, however, to this rule which is that the algorithm also updates homologue connections between the same regions in either hemisphere, given that tractography is known to be less accurate when accounting for this connectivity.

For the Stuart-Landau model, we used *α* = *ς* = 0.0004 and continue until the algorithm converges. For each iteration we compute the model results as the average over as many simulations as there are participants. The resulting ***C*** coupling matrix is the GEC.

### Framework for Quantum-like Analysis

#### Superposition and Interference States: Hilbert Space

Interestingly, recent advances in physics have shown so-called QL interference at the macroscopic level in systems of non-quantum (classical) coupled oscillators (Scholes, 2024, 2025; Scholes and Amati, 2025), which can be conceptualised within the Växjö interpretation of quantum mechanics (Khrennikov, 2010). Similar to a real quantum state, interactions between different QL networks of oscillators are able to produce interference superposition.

QL states can be expressed as vectors in a Hilbert space and any interference effect can thus be described in the same way. Briefly, a Hilbert space ℋis a complete vector space over a field ℤ. Here we use a real field with the inner product ⟨*x, y*⟩, thereby generalising the Cartesian dot product. So, for any vectors *x, y, z* in ℋ and with scalars *α* ∈ ℤ, the following equalities are satisfied:

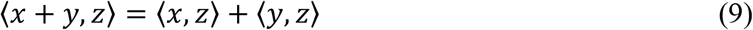

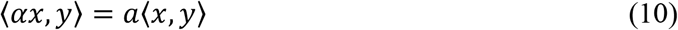

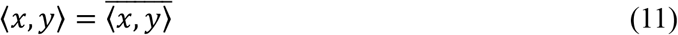

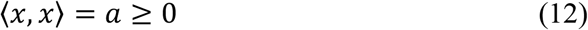

where overbar signifies the complex conjugate. The norm is defined in terms of the inner product: 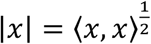.

#### Definition of Spectral Gaps

The spectral gap of a connectivity matrix can characterise the potential emergent states. In whole-brain modelling, the spectral gap properties of the GEC shape the emergence of multiple dynamical states. In our framework, these correspond to cluster-synchronisation configurations that are intrinsically metastable and collectively generate a rich dynamical repertoire. Larger spectral gaps favour the formation of these metastable interference states and thus provide a quantitative proxy for the degree of QL behaviour in the system.

Following the optimisation of the GEC in a whole-brain model, we performed an eigen decomposition of the GEC. The spectral gaps were derived from the distribution of the eigenvalues. Let the eigenvalues of the GEC coupling be sorted in descending order *λ*_0_ ≥ *λ*_1_ ≥ … ≥ *λ*_*n*−1_. We calculated the distribution of absolute spacings between consecutive eigenvalues. To identify significant gaps in the spectrum, we defined a threshold given by the mean of the spacings plus one standard deviation of the mean. The measurement of spectral gaps was calculated as the sum of all spacings exceeding this threshold and is thus a measure of the structural entanglement.

#### Turbulent Vortex Space: Spatiotemporal Metastability

As has been shown, turbulence is not restricted to fluid dynamics but is also found in other physical systems including coupled oscillators (Kawamura *et al*., 2007) and brains (Deco and Kringelbach, 2020; Deco *et al*., 2023; Deco *et al*., 2025c). Yoshiki Kuramoto used the theory of coupled oscillators to show turbulence in fluid dynamics (Kuramoto, 1984), suggestive of how turbulence could be important not only for energy transfer but for efficient information transfer. Specifically, this framework defines the Kuramoto local order parameter, representing a spatial average of the complex phase factor of the local oscillators weighted by the coupling. Thus, the Kuramoto local order parameter gives the level of synchronisation of the local phases around a specific point, which is here called ‘Kuramoto vorticity’. Importantly, the level of amplitude turbulence is defined as the standard deviation of the modulus of ‘Kuramoto vorticity’ and can be applied to the empirical data of any physical system. Remarkably, using this measure, the human brain was found to exhibit a similar turbulent power law, strongly suggesting the presence of a cascade of efficient information processing across scales (Deco and Kringelbach, 2020; Deco *et al*., 2023; Deco *et al*., 2025c).

We measure turbulence by first defining the *Kuramoto local order parameter* and then taking the standard deviation of the modulus across time and space (Kawamura *et al*., 2007). The local Kuramoto order parameter *R* is thus computed by

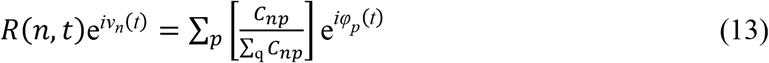

where *φ*_*p*_(*t*) are the phases of the spatiotemporal data, *ν*_*n*_(*t*) are the phases of the Kuramoto order parameter and *C*_*np*_ is the local neighbourhood weighting exponential kernel between brain regions *n* and *p* given by

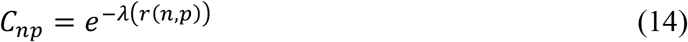

Where λ defines the spatial scaling and *r*(*n, p*) is the Euclidean distance between the brain regions *n* and *p*. Here, we used a λ=0.18 mm^-1^ obtained after fitting the anatomical exponential-rule to the dMRI tractography (Deco and Kringelbach, 2020). Thus, the Turbulence, *T*, is defined as the standard deviation of the local Kuramoto order parameter across space and time:

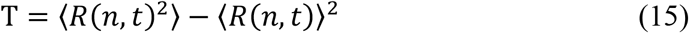

where the brackets ⟨ ⟩ denotes average across vertex space and time. Note, that the local Kuramoto order parameter *R*(*n, t*) in fact defines a new spatiotemporal dynamic in the so-called vortex space.

Importantly, an analytical equation can be found for the evolution of the local Kuramoto order parameters, describing the dynamics of the vortex space, defined in a partition of the original system, here in the Schaefer1000 parcellation (*M* = 1000) (Deco *et al*., 2025a). In this case, the local Kuramoto order parameter is defined in a partitioned Schaefer100 (*N*=100) as follows:

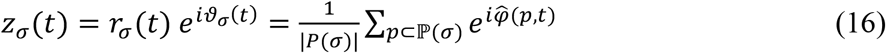

This uses a partition ℘ = (ℙ(1), … ., ℙ(*N*)) of the index set [1, …, *M*] by the Schaefer parcellation *N*=100 of the original system defined in the Schaefer parcellation. Note that the *z*_σ_ (*t*) expresses the vorticity in the partitioned space (“partitioned Local Kuramoto order parameters”) by averaging spatially the complex phase factor of the local phases 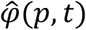 from the BOLD time series signals in the fine Schaefer 1000 parcellation. The partitioned vorticity, *z*_*σ*_ (*t*) reflects the level of synchronisation of the local phases in a given partition *σ* (here in the Schaefer100 parcellation) and is defining a partitioned version of the vortex space.

#### Quantum-like Entanglement in Vortex Space: The Växjö Interference

To perform probabilistic interference analysis, the partitioned vorticity *z*_*σ*_ (*t*) was discretised, binning it in five states *S*_*i*_ = [1,2,3,4,5]. This transformation allows for the definition of a discrete probability space where the state of a brain region *σ* in the Schaefer parcellation100 at any time *t* is categorised by its level of local collective oscillation.

In a classical stochastic system, the interaction between two regions *i* and *j* should obey the Law of Total Probability (LTP). The LTP posits that the marginal probability of region *i* being in a specific state *m* can be perfectly reconstructed by summing the conditional probabilities of region *i* across all possible states *k* of region *j*, i.e.

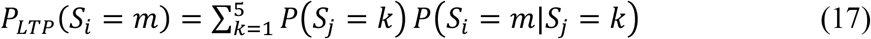

Any significant deviation from this identity indicates a “contextual” or “quantum-like” interaction, where the presence of region *j* creates an interference effect that biases the probability distribution of region *i* in a non-additive manner. Following the Växjö framework for contextual probability, we defined the Växjö Interference Term *δ*_*ij*_ as the normalised violation of the LTP for the pair (*i, j*). First, we calculated the absolute deviation (the “interference residue”) for each state *m*:

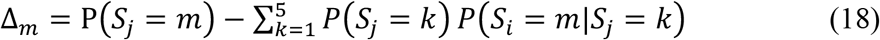

To account for the probabilistic weight of the interaction, the final interference term *δ*_*ij*_ was computed by normalising the residue by the geometric mean of the joint probabilities across all states:

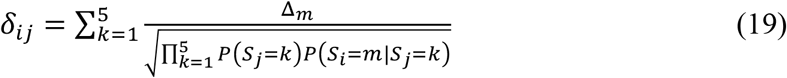

VIC (Växjö Interference Connectivity) is the matrix of *δ*_*ij*_ elements. In figures, VIC is calculated as the grand mean of *δ*_*ij*_ across all pairs, excluding cases where the denominator approached zero (infinite interference).

### Modified Connectome-based Prediction Modelling (mCPM)

We used a modified CPM framework (Greene *et al*., 2018; Shen *et al*., 2017) to predict one-year symptom change for held-out participants from their connectivity features in the BANDA cohort of 150 participants stratified into Control (CNT), Anxious (ANX), and Depressed (DEP) groups. The target behavioural variable was RCADS, which provides a quantitative measure of emotional symptom severity. To evaluate both concurrent and prospective predictive validity, we defined two distinct targets: 1) *Baseline symptoms*: RCADS total score at the time of the resting-state fMRI scan, and 2) *One-year follow-up symptoms*: RCADS total score at a one-year clinical follow-up.

To strictly prevent data leakage during prospective prediction, the initial ranking of feature relevance was computed using Pearson correlations between the connectivity features and the baseline symptom scores. This isolates the prospective target from feature selection. The target variable for the model—the one-year follow-up symptom scores—was completely isolated from this feature ranking step. The optimal correlation threshold for feature inclusion was then dynamically selected within the inner loops of the nested cross-validation using the outer training data.

Thus, to reduce the dimensionality of the feature space relative to the sample size, we pre-selected features on the basis of their association with baseline symptoms. We computed the Pearson correlation with baseline symptoms, producing a vector of correlations that guided selection throughout the procedure. A correlation threshold controlled what entered the regression at each step. Candidate thresholds ranged from 0.08 to the maximum absolute correlation minus 0.03 in steps of 0.01.

We predicted one-year follow-up symptoms for held-out participants from their connectivity features using ridge regression. Ridge was chosen because of the strong collinearity inherent in whole-brain connectivity matrices, where many edges share variance and an L1 penalty would be unstable. Model performance was assessed through a nested cross-validation scheme designed to select the feature threshold and the regularisation parameter without reusing the outer test data for either.

The outer loop consisted of ten-fold cross-validation on one-year follow-up scores. Within each outer training fold, we ran a 5-fold inner cross-validation across the candidate thresholds. For every candidate threshold, the selected feature subset was standardised using the inner training mean and standard deviation, and entered into a ridge regression fitted through a three-fold internal cross-validation over the regularisation path. The threshold maximising the Pearson correlation between observed and predicted inner-validation scores was retained for that outer fold.

The selected threshold was then applied to the full outer training set. Features were re-standardised using outer training statistics and a ridge regression was fitted, with the regularisation parameter chosen by five-fold internal cross-validation. Held-out outer test participants were projected into the outer training standardised space and their one-year follow-up values predicted from the fitted model. For each repetition, predictive performance was quantified using two complementary metrics computed across all participants: i) Pearson correlation between observed and predicted behavioural scores; ii) Cross-validated coefficient of determination (*R*^2^), computed as:

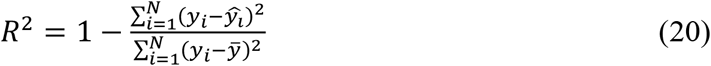

To localise the brain regions most critical for clinical prediction, we performed a post-hoc rendering. We identified the predictive sub-network (edges selected in the filtering stage) and calculated for each brain region the averaged Växjö interference with all the other regions in the brain. This allowed to localise the specific functional nodes where Växjö interference effects were most strongly predicting the RCADS symptom groups.

To evaluate the HCP dataset, we utilised an inference-based mCPM framework to model the g-factor and individual cognitive performance across four domains: *Card Sorting, Picture Vocabulary, Progressive Matrices* (PMAT24) and *Processing Speed*. Unlike the prospective prediction pipeline used for BANDA, this inference framework focuses on characterising population-level neural masks. Specifically, feature selection was performed across the entire dataset to identify features with the strongest global association to the behavioural target. Once the neural mask was defined, a ridge regression was fitted using a 10-fold cross-validation scheme (repeated 100 times) to evaluate the robustness and stability of the multivariate mapping rather than its out-of-sample predictive generalisability. In both cases, two types of inference models were constructed separately using QL interference VIC features and conventional FC features, enabling a direct comparison between the two representations.

### The cost of Cognition: Brain Computability

Computability is defined as the efficiency of a brain’s functional reconfiguration. It quantifies the “accuracy” of a transition from a baseline resting-state to a task-active state relative to the “biological effort” (*cost*) required to manifest that change. For each participant and task, the computability is calculated as the correlation between the observed log-ratio of GEC connectivity changes in each task relative to the whole-brain model achieved GEC task based on the GEC rest modulated by the neuroreceptors (specified below). The target GEC task was calculated for the top 10% of participants scoring highly on intelligence (as indexed by the g-factor), since there are assumed to perform more optimal computation. Thus, this measure characterises how successfully the brain has reconfigured. We normalise this measure by the cost computed as the pressure required by the modulatory system to perform the reconfiguration. This means that a high *computability index* identifies a system capable of precise, high-fidelity reconfigurations for a relatively low biological price.

#### Neuroreceptor Mapping and Normalisation

Receptor densities were assessed using PET tracer studies for 19 different receptors and transporters across nine neurotransmitter systems from more than 1,200 healthy participants, based on data provided by Hansen and colleagues (Hansen *et al*., 2022). These neurotransmitter systems include dopamine (D1, D2, DAT, noradrenaline (NET), serotonin (5HT1A, 5HT1B, 5HT2A, 5HT4, 5HT6, 5HTT), acetylcholine (A4B2, M1, VAChT), glutamate (mGLUR5, NMDA), GABA (GABA-A), histamine (H3), cannabinoid (CB1), and opioid (MOR). The volumetric PET images were aligned to the MNI-ICBM 152 nonlinear 2009 (version c, asymmetric) template, averaged across participants within each study, and then parcellated. For receptors/transporters with multiple mean images from the same tracer (5HT1B, D2, VAChT), a weighted average was applied.

Importantly, the neurotransmitter maps were normalised following the procedure of Fulcher and Fornito (Fulcher and Fornito, 2016). The maps are z-scored across the 100 regions in the Schaefer100 parcellation and then non-linearly normalised by a sigmoid function to reduce the influence of outliers:

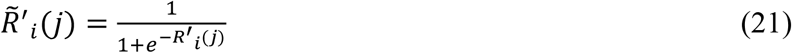

where *R*^′^_*i*_(*j*) signifies the receptor *j* in the region *i* from the original neurotransmitter maps provided by Hansen and colleagues (Hansen *et al*., 2022).

Hence, the normalised neurotransmitter maps are given by

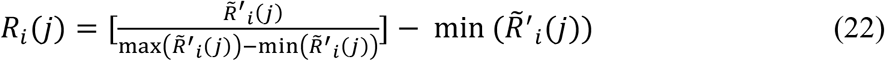

#### Neuromodulation in the Whole-brain Model: NEMO in Generative Space

Here, we improved our computability approach previously published as Neuromodulation in the Whole-brain Model (NEMO) (Deco *et al*., 2026). We define computability as the efficiency with which a cognitive state (task) can be reached from a baseline state (rest) through a transformation constrained by the molecular (receptor) landscape. In this paper, we apply the same philosophy as NEMO but in generative space. Specifically, we measure the effect of the neuromodulation in generative space, i.e., we measure computability by the level of fitting of the neuromodulated model of a specific task in terms of the underlying GEC matrices. We fit the log ratio of the GEC elements (empirical versus neuromodulated model). We use a bilinear Ordinary Least squares (OLS) framework. In the whole-brain model, the transition from the resting-state (*C*_*rest*_) to a task-active state (*C*_*task*_) is modelled as a reconfiguration driven by an edge modification of the GEC with a weighted sum of the normalised receptor densities at the extreme of each edge as specified below. We define the regression target as the log-ratio between the empirical task-GEC between and the empirical rest-GEC. For each element of the connectivity matrix we optimized

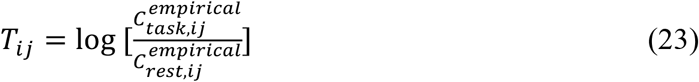

The use of a log-ratio captures the relative scaling (potentiation or inhibition) of directed edges. We construct a high-dimensional design matrix for all off-diagonal edges to solve for the weights using Ordinary Least Squares (OLS). The model includes two primary modulations for each edge of the model GEC: 1) A weighted sum over all neuroreceptor, where each summand is modulated by the product of the each receptor *k* density at node *i* and node *j* (i.e. *R*_*i*_(*k*)*R*_*j*_(*k*)), representing simple co-expression facilitation, 2) Interaction Effects: The synergistic product of two different receptors, *k* and *l* given by *R*_*i*_(*k*)*R*_*j*_(*k*)*R*_*i*_(*l*)*R*_*j*_(*l*) across the two nodes, representing cross-system neuromodulation. In mathematical terms:

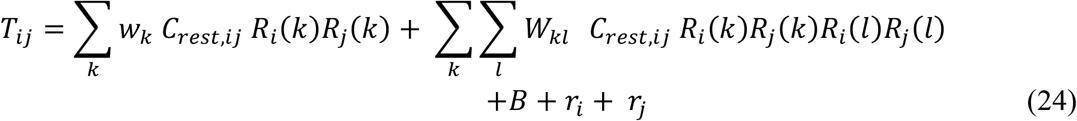

Please note that while the neuroreceptor densities capture the molecular drivers of the rest-to-task transition, we incorporated a global bias to account for uniform, brain-wide shifts in average connectivity strength, and a regional offset to control for node-specific factors—such as local vascular biases, intrinsic excitability, or topological hubness—that influence an area’s capacity for reconfiguration regardless of its specific neuroreceptor profile. To sum up, the transition from a resting-state configuration to a task-optimal state is modelled as a parsimonious reorganisation constrained by the neuroreceptor landscape.

We calculated the computability for each participant as the average of the computability for each of the seven HCP cognitive tasks. We define the total transition cost (**K**) as the L1-norm of the parameters required to reconstruct the observed change in Effective Connectivity. By summing the absolute weights of the linear model, we quantify the total “pressure” applied to the system to effect the transition. Mathematically, for each participant and task, the cost is defined as:

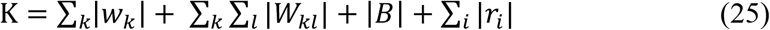

For each participant, the final cost is the sum of the cost over all tasks. This definition assumes that a “low-cost” transition is one where the brain utilises its existing molecular architecture efficiently, whereas a “high-cost” transition requires significant regional deviations or heavy reliance on complex receptor interactions.

Once the weights were optimized, we evaluated the model’s performance by reconstructing the predicted (simulated) task-specific effective connectivity matrix (*C*_*task*_) from the model-optimised *C*_*rest*_. This was achieved by applying the exponential of the predicted neuromodulated reconfiguration to the resting-state baseline:

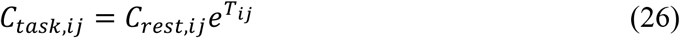

Where *T*_*ij*_ is the predicted log-ratio matrix reconstructed from the neuroreceptor maps. The fitness of the model was defined as the accuracy with which the neuromodulated model could reconstruct the empirical elements of the task GEC. We calculated the Pearson correlation coefficient between the vectorised off-diagonal elements of the empirical task connectivity and the model-predicted task connectivity and to ensure a normal distribution for statistical comparison across groups, we applied the Fisher z-transformation to this correlation. This z-score represents the reconfiguration fitness, quantifying how much of the actual task-driven connectivity organisation is successfully captured by the neuroreceptor-constrained model. Finally, we defined computability as the efficiency of the reconfiguration, calculated as the ratio between the fitness (accuracy) and the cost (effort). This metric provides a formal measure of “computational ease”. A higher index indicates that the task-specific GEC is highly computable from the resting-state given the brain’s molecular and regional constraints, achieving high reconstruction fidelity with minimal parametric cost.

### Energy Dissipation and Information Processing

We also computed the energy consumed by each participant, in the neuromodulated reconfiguration necessary to achieve each of the seven given tasks. We were able to compute this by applying elements of stochastic thermodynamics. Non-equilibrium systems are central to many scientific fields, but fundamental challenges remain, especially in defining microscopic entropy production— a key measure of irreversibility. In thermodynamic terms, the change of entropy Δ*S* for a reversible process is given by:

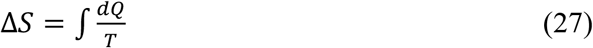

where *dQ* is the heat exchanged and *T* is temperature. For irreversible processes,

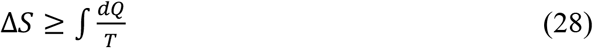

and the difference defines entropy production *H*_*P*_, i.e.:

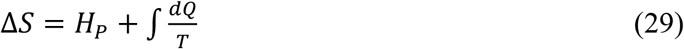

The rates of change derived with respect to time (with the dot on the top of a function signifying the time derivative) are given by:

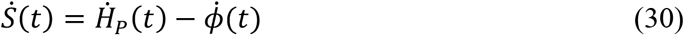

In this equation 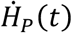 is the entropy production rate of the system, which (according to the second law of thermodynamics) is always non-negative, and 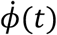 is the entropy flux rate from the system to the environment. For systems in a non-equilibrium steady-state, 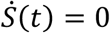 which implies 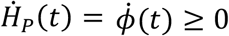. Please note that it is only for thermodynamic equilibrium that the equality in this equation is expected to hold.

In the brain, 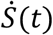, the rate of change of entropy, can be interpreted as ‘computation’, that is the flexible mapping of input with output through internal transformations and consequently as a measure of information processing. On the other hand, the 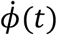 is the entropy flux can be interpreted as the energy consumption for performing a particular computation, and 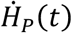 is the level of non-equilibrium. These fundamental thermodynamic quantities can be calculated analytically when the system is a linear Langevin system, which is exactly the case for our linearised Stuart-Landau model. Let us consider the following Langevin system using a simplifying notation of *δ****u***= ***x***:

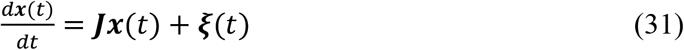

The ***ξ***(*t*) are 2N independent standard Wiener processes with covariance:

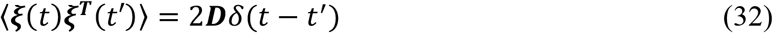

Therefore ***D*** is a 2*M* × 2*M* diagonal matrix (but now generalised to the possibility that the diagonal elements could be different). The angular brackets indicate the mathematical expectation over time and the *δ*(*t*) is the Dirac’s delta function.

Given this, it is straightforward to derive the temporal evolution of the covariance matrix, ***θ*** = ⟨***x***(*t*)***x***^***T***^(*t*′)⟩ − ⟨***x***(*t*)⟩⟨***x***(*t*′)⟩, which is given by:

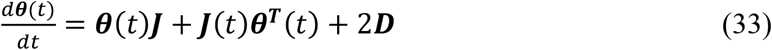

The probability distribution *P*(***x***, *t*) associated with the Langevin equation is described by following Fokker-Planck equation:

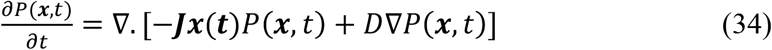

Where ∇ denotes the spatial derivative with respect to ***x***. The above Fokker-Planck equation can be expressed as the following continuity equation:

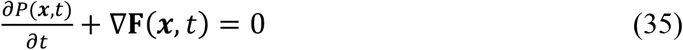

where the probability current is given by:

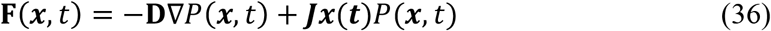

The stationary solution can be derived analytically and is given by following multivariate Gaussian distribution:

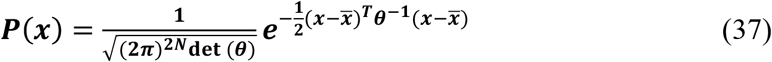

The entropy is defined by:

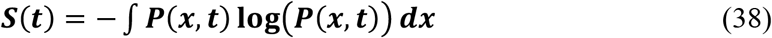

As shown by Landi and colleagues (Landi *et al*., 2013) and Gilson and colleagues (Gilson *et al*., 2023), the rate of change of entropy can be decomposed as:

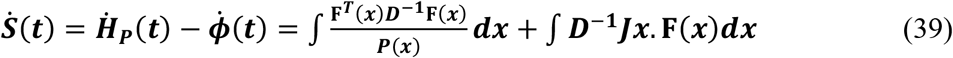

Where the stationary probability current **F**(***x***) is given by

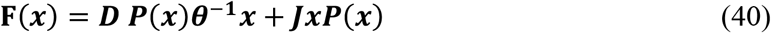

From these equations, following the lead of Landi and colleagues, we can express the information processing (as the rate of the change in entropy):

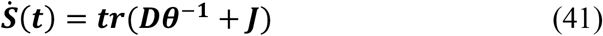

while the entropy production can be expressed as:

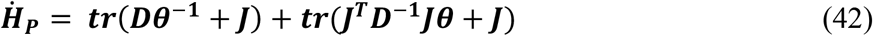

and energy consumption (measure as entropy flow out of the system) as follows:

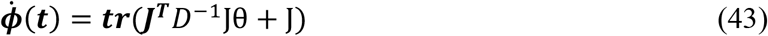

where tr is the trace of the matrix.

We use Equation (43) for computing the energy consumption of each participant in switching from resting to a task (averaged over all 7 tasks). We do this, as specified in the main body of the paper, for the full trained GEC (i.e. the QL model) and for the poorly trained GEC (i.e. the non-QL model). We initialised the iteration of Equation (43) with the one corresponding to the participant specific resting model, and for further iterations we switched to the corresponding specific task model and averaged over all tasks.

## References

Balasubramanian, V. (2021) Brain power. Proc. Natl. Acad. Sci. U.S.A. 118, e2107022118.

Barch, D. M., Burgess, G. C., Harms, M. P., Petersen, S. E., Schlaggar, B. L., Corbetta, M., Glasser, M. F., Curtiss, S., Dixit, S., Feldt, C., Nolan, D., Bryant, E., Hartley, T., Footer, O., Bjork, J. M., Poldrack, R., Smith, S., Johansen-Berg, H., Snyder, A. Z., Van Essen, D. C. and Consortium, W. U.-M. H. (2013) Function in the human connectome: task-fMRI and individual differences in behavior. NeuroImage 80, 169–189.

Bulinski, A. and Khrennikov, A. (2004) Nonclassical total probability formula and quantum interference of probabilities. Statistics & Probability Letters 70, 49–58.

Deco, G. and Kringelbach, M. L. (2020) Turbulent-like dynamics in the human brain. Cell Reports 33, 108471.

Deco, G. and Kringelbach, M. L. (2025) Whole-brain modelling. Cartography of the dynamics of Mind. Oxford University Press: Oxford.

Deco, G., Kringelbach, M. L., Jirsa, V. and Ritter, P. (2017) The Dynamics of Resting Fluctuations in the Brain: Metastability and its Dynamical Core [bioRxiv 065284]. Sci. Rep. 7, 3095.

Deco, G., Leibana Garcia, S., Sanz Perl, Y., Tagliazucchi, E. and Kringelbach, M. L. (2023) The effect of turbulence in brain dynamics information transfer measured with magnetoencephalography. Commun Phys 6, 74.

Deco, G., Perl, Y. S., Feng, J. and Kringelbach, M. L. (2025a) The turbulent brain: Modelling vortex interactions for understanding human cognition. Network neuroscience.

Deco, G., Perl, Y. S., Greenstein, N., Chandaria, S., Scholes, G. and Kringelbach, M. L. (2025b) Quantum-like dynamics in the human brain. bioRxiv, 2025.2010.2002.680057.

Deco, G., Perl, Y. S., Jerotic, K., Escrichs, A. and Kringelbach, M. L. (2025c) Turbulence as a framework for brain dynamics in health and disease. Neurosci Biobehav Rev 169, 105988.

Deco, G., Ponce-Alvarez, A., Hagmann, P., Romani, G. L., Mantini, D. and Corbetta, M. (2014) How local excitation-inhibition ratio impacts the whole brain dynamics. J. Neurosci. 34, 7886– 7898.

Deco, G., Sanz Perl, Y., Johnson, S., Bourke, N., Carhart-Harris, R. L. and Kringelbach, M. L. (2024) Different hierarchical reconfigurations in the brain by psilocybin and escitalopram for depression. Nature Mental Health 2, 1096–1110.

Deco, G., Sanz Perl, Y., Luppi, A., Gini, S., Gozzi, A., Carhart-Harris, R., Chandaria, S. and Kringelbach, M. L. (2025d) The cost of cognition: Measuring the energy consumption of nonequilibrium computation. bioRxiv, 2025.2006.2018.660368.

Deco, G., Sanz Perl, Y., Tagliazucchi, E. and Kringelbach, M. L. (2022) The INSIDEOUT framework provides precise signatures of the balance of intrinsic and extrinsic dynamics in brain states. Commun. Biol. 5, 572.

Deco, G., Sanz Perl, Y., Vohryzek, J., Luppi, A. I. and Kringelbach, M. L. (2026) Neurotransmission-modulated whole-brain computation captures full task repertoire. Cell Rep 45, 116816.

Deco, G., Vidaurre, D. and Kringelbach, M. L. (2021) Revisiting the global workspace orchestrating the hierarchical organization of the human brain. Nat. Human Behav. 5, 497–511.

Dubois, J., Galdi, P., Paul, L. K. and Adolphs, R. (2018) A distributed brain network predicts general intelligence from resting-state human neuroimaging data. Philos. Trans. R. Soc. Lond. B. Biol. Sci. 373.

Escrichs, A., Sanz Perl, Y., Fisher, P. M., Martinez-Molina, N., E, G. G., Frokjaer, V. G., Kringelbach, M. L., Knudsen, G. M. and Deco, G. (2024) Whole-brain turbulent dynamics predict responsiveness to pharmacological treatment in major depressive disorder. Mol Psychiatry 30, 1069–1079.

Escrichs, A., Sanz Perl, Y., Uribe, C., Camara, E., Turker, B., Pyatigorskaya, N., Lopez-Gonzalez, A., Pallavicini, C., Panda, R., Annen, J., Gosseries, O., Laureys, S., Naccache, L., Sitt, J. D., Laufs, H., Tagliazucchi, E., Kringelbach, M. L. and Deco, G. (2022) Unifying turbulent dynamics framework distinguishes different brain states. Commun. Biol. 5, 638.

Fulcher, B. D. and Fornito, A. (2016) A transcriptional signature of hub connectivity in the mouse connectome. Proc. Natl. Acad. Sci. U.S.A. 113, 1435–1440.

Gilson, M., Deco, G., Friston, K. J., Hagmann, P., Mantini, D., Betti, V., Romani, G. L. and Corbetta, M. (2017) Effective connectivity inferred from fMRI transition dynamics during movie viewing points to a balanced reconfiguration of cortical interactions. NeuroImage 180, 534– 546.

Gilson, M., Tagliazucchi, E. and Cofré, R. (2023) Entropy production of multivariate Ornstein-Uhlenbeck processes correlates with consciousness levels in the human brain. Physical Review E 107, 024121.

Glasser, M. F., Sotiropoulos, S. N., Wilson, J. A., Coalson, T. S., Fischl, B., Andersson, J. L., Xu, J., Jbabdi, S., Webster, M., Polimeni, J. R., Van Essen, D. C., Jenkinson, M. and Consortium, W. U.-M. H. (2013) The minimal preprocessing pipelines for the Human Connectome Project. NeuroImage 80, 105–124.

Greene, A. S., Gao, S., Scheinost, D. and Constable, R. T. (2018) Task-induced brain state manipulation improves prediction of individual traits. Nat. Commun. 9, 2807.

Griffanti, L., Salimi-Khorshidi, G., Beckmann, C. F., Auerbach, E. J., Douaud, G., Sexton, C. E., Zsoldos, E., Ebmeier, K. P., Filippini, N., Mackay, C. E., Moeller, S., Xu, J., Yacoub, E., Baselli, G., Ugurbil, K., Miller, K. L. and Smith, S. M. (2014) ICA-based artefact removal and accelerated fMRI acquisition for improved resting state network imaging. NeuroImage 95, 232–247.

Hansen, J. Y., Shafiei, G., Markello, R. D., Smart, K., Cox, S. M. L., Norgaard, M., Beliveau, V., Wu, Y., Gallezot, J. D., Aumont, E., Servaes, S., Scala, S. G., DuBois, J. M., Wainstein, G., Bezgin, G., Funck, T., Schmitz, T. W., Spreng, R. N., Galovic, M., Koepp, M. J., Duncan, J. S., Coles, J. P., Fryer, T. D., Aigbirhio, F. I., McGinnity, C. J., Hammers, A., Soucy, J. P., Baillet, S., Guimond, S., Hietala, J., Bedard, M. A., Leyton, M., Kobayashi, E., Rosa-Neto, P., Ganz, M., Knudsen, G. M., Palomero-Gallagher, N., Shine, J. M., Carson, R. E., Tuominen, L., Dagher, A. and Misic, B. (2022) Mapping neurotransmitter systems to the structural and functional organization of the human neocortex. Nat. Neurosci. 25, 1569–1581.

Hausfather, Z., Drake, H. F., Abbott, T. and Schmidt, G. A. (2020) Evaluating the Performance of Past Climate Model Projections. Geophysical Research Letters 47, e2019GL085378.

Hubbard, N. A., Bauer, C. C. C., Siless, V., Auerbach, R. P., Elam, J. S., Frosch, I. R., Henin, A., Hofmann, S. G., Hodge, M. R., Jones, R., Lenzini, P., Lo, N., Park, A. T., Pizzagalli, D. A., Vaz-DeSouza, F., Gabrieli, J. D. E., Whitfield-Gabrieli, S., Yendiki, A. and Ghosh, S. S. (2024) The Human Connectome Project of adolescent anxiety and depression dataset. Scientific data 11, 837.

Hubbard, N. A., Siless, V., Frosch, I. R., Goncalves, M., Lo, N., Wang, J., Bauer, C. C. C., Conroy, K., Cosby, E., Hay, A., Jones, R., Pinaire, M., Vaz De Souza, F., Vergara, G., Ghosh, S., Henin, A., Hirshfeld-Becker, D. R., Hofmann, S. G., Rosso, I. M., Auerbach, R. P., Pizzagalli, D. A., Yendiki, A., Gabrieli, J. D. E. and Whitfield-Gabrieli, S. (2020) Brain function and clinical characterization in the Boston adolescent neuroimaging of depression and anxiety study. NeuroImage. Clinical 27, 102240.

Kaiser, R. H., Andrews-Hanna, J. R., Wager, T. D. and Pizzagalli, D. A. (2015) Large-Scale Network Dysfunction in Major Depressive Disorder: A Meta-analysis of Resting-State Functional Connectivity. JAMA psychiatry 72, 603–611.

Kawamura, Y., Nakao, H. and Kuramoto, Y. (2007) Noise-induced turbulence in nonlocally coupled oscillators. Physical review. E, Statistical, nonlinear, and soft matter physics 75, 036209.

Kessler, R. C., Berglund, P., Demler, O., Jin, R., Merikangas, K. R. and Walters, E. E. (2005) Lifetime prevalence and age-of-onset distributions of DSM-IV disorders in the National Comorbidity Survey Replication. Archives of general psychiatry 62, 593–602.

Khera, A. V., Chaffin, M., Aragam, K. G., Haas, M. E., Roselli, C., Choi, S. H., Natarajan, P., Lander, E. S., Lubitz, S. A., Ellinor, P. T. and Kathiresan, S. (2018) Genome-wide polygenic scores for common diseases identify individuals with risk equivalent to monogenic mutations. Nature genetics 50, 1219–1224.

Khrennikov, A. (2010) Ubiquitous quantum structure. Springer.

Khrennikov, A. Y. (2021) Contextual approach to quantum formalism. Springer: New York.

Kringelbach, M. L. (2005) The orbitofrontal cortex: linking reward to hedonic experience. Nature Reviews Neuroscience 6, 691–702.

Kringelbach, M. L. and Deco, G. (2020) Brain States and Transitions: Insights from Computational Neuroscience. Cell Reports 32, 108128.

Kringelbach, M. L. and Deco, G. (2026) One operator to rule them all: Unifying connectome harmonics, turbulence and complex harmonics in brain dynamics. bioRxiv, 2026.2006.2005.730423.

Kringelbach, M. L., Rosas, F., Laukkonen, R., Chandaria, S., Sanz Perl, Y. and Deco, G. (2026) The Entangled Loop: Emotion, quantum-like interference and hierarchy in the architecture of consciousness. PsyArXiv, osf.io/preprints/psyarxiv/besw9_v2.

Kringelbach, M. L., Sanz Perl, Y. and Deco, G. (2024) The Thermodynamics of Mind. Trends Cogn Sci 28, 568–581.

Kringelbach, M. L., Sanz Perl, Y., Tagliazucchi, E. and Deco, G. (2023) Toward naturalistic neuroscience: Mechanisms underlying the flattening of brain hierarchy in movie-watching compared to rest and task. Science Advances 9, eade6049.

Kuramoto, Y. (1984) Chemical Oscillations,Waves, and Turbulence. Springer-Verlag: Berlin.

Landi, G. T., Tomé, T. and de Oliveira, M. J. (2013) Entropy production in linear Langevin systems. Journal of Physics A: Mathematical and Theoretical 46, 395001.

Levy, W. B. and Calvert, V. G. (2021) Communication consumes 35 times more energy than computation in the human cortex, but both costs are needed to predict synapse number. Proc. Natl. Acad. Sci. U.S.A. 118, e2008173118.

Marek, S., Tervo-Clemmens, B., Calabro, F. J., Montez, D. F., Kay, B. P., Hatoum, A. S., Donohue, M. R., Foran, W., Miller, R. L., Hendrickson, T. J., Malone, S. M., Kandala, S., Feczko, E., Miranda-Dominguez, O., Graham, A. M., Earl, E. A., Perrone, A. J., Cordova, M., Doyle, O., Moore, L. A., Conan, G. M., Uriarte, J., Snider, K., Lynch, B. J., Wilgenbusch, J. C., Pengo, T., Tam, A., Chen, J., Newbold, D. J., Zheng, A., Seider, N. A., Van, A. N., Metoki, A., Chauvin, R. J., Laumann, T. O., Greene, D. J., Petersen, S. E., Garavan, H., Thompson, W. K., Nichols, T. E., Yeo, B. T. T., Barch, D. M., Luna, B., Fair, D. A. and Dosenbach, N. U. F. (2022) Reproducible brain-wide association studies require thousands of individuals. Nature 603, 654–660.

Martinez-Molina, N., Sanz-Perl, Y., Escrichs, A., Kringelbach, M. L. and Deco, G. (2024) Turbulent dynamics and whole-brain modeling: toward new clinical applications for traumatic brain injury. Front Neuroinform 18, 1382372.

McGorry, P. D. and Mei, C. (2018) Early intervention in youth mental health: progress and future directions. Evidence Based Mental Health 21.

Morfini, F., Kucyi, A., Zhang, J., Bauer, C. C. C., Bloom, P. A., Pagliaccio, D., Hubbard, N. A., Rosso, I. M., Yendiki, A., Ghosh, S. S., Pizzagalli, D. A., Gabrieli, J. D. E., Whitfield-Gabrieli, S. and Auerbach, R. P. (2025) Brain functional connectivity predicts depression and anxiety during childhood and adolescence: A connectome-based predictive modeling approach. Imaging Neurosci (Camb) 3.

Navarro Schroder, T., Haak, K. V., Zaragoza Jimenez, N. I., Beckmann, C. F. and Doeller, C. F. (2015) Functional topography of the human entorhinal cortex. eLife 4, e06738.

Oostenveld, R., Fries, P., Maris, E. and Schoffelen, J. M. (2011) FieldTrip: Open source software for advanced analysis of MEG, EEG, and invasive electrophysiological data. Comput. Intell. Neurosci. 2011, 156869.

Paus, T., Keshavan, M. and Giedd, J. N. (2008) Why do many psychiatric disorders emerge during adolescence? Nat. Rev. Neurosci. 9, 947–957.

Pizzagalli, D. A. (2022) Toward a better understanding of the mechanisms and pathophysiology of anhedonia: are we ready for translation? American Journal of Psychiatry 179, 458–469.

Ponce-Alvarez, A. and Deco, G. (2024) The Hopf whole-brain model and its linear approximation. Sci. Rep. 14, 2615.

Pope, M., Seguin, C., Varley, T. F., Faskowitz, J. and Sporns, O. (2023) Co-evolving dynamics and topology in a coupled oscillator model of resting brain function. NeuroImage 277, 120266.

Rapee, R. M., Oar, E. L., Johnco, C. J., Forbes, M. K., Fardouly, J., Magson, N. R. and Richardson, C. E. (2019) Adolescent development and risk for the onset of social-emotional disorders: A review and conceptual model. Behav Res Ther 123, 103501.

Salimi-Khorshidi, G., Douaud, G., Beckmann, C. F., Glasser, M. F., Griffanti, L. and Smith, S. M. (2014) Automatic denoising of functional MRI data: combining independent component analysis and hierarchical fusion of classifiers. NeuroImage 90, 449–468.

Sawyer, S. M., Azzopardi, P. S., Wickremarathne, D. and Patton, G. C. (2018) The age of adolescence. The lancet child & adolescent health 2, 223–228.

Schaefer, A., Kong, R., Gordon, E. M., Laumann, T. O., Zuo, X. N., Holmes, A. J., Eickhoff, S. B. and Yeo, B. T. T. (2018) Local-Global Parcellation of the Human Cerebral Cortex from Intrinsic Functional Connectivity MRI. Cereb. Cortex 28, 3095–3114.

Scholes, G. D. (2024) Quantum-like states on complex synchronized networks. Proceedings of the Royal Society A 480, 20240209.

Scholes, G. D. (2025) Quantum-like states from classical systems. arXiv, 2507.00967

Scholes, G. D. and Amati, G. (2025) Quantumlike Product States Constructed from Classical Networks. Physical review letters 134, 060202.

Setsompop, K., Kimmlingen, R., Eberlein, E., Witzel, T., Cohen-Adad, J., McNab, J. A., Keil, B., Tisdall, M. D., Hoecht, P., Dietz, P., Cauley, S. F., Tountcheva, V., Matschl, V., Lenz, V. H., Heberlein, K., Potthast, A., Thein, H., Van Horn, J., Toga, A., Schmitt, F., Lehne, D., Rosen, B. R., Wedeen, V. and Wald, L. L. (2013) Pushing the limits of in vivo diffusion MRI for the Human Connectome Project. NeuroImage 80, 220–233.

Shen, X., Finn, E. S., Scheinost, D., Rosenberg, M. D., Chun, M. M., Papademetris, X. and Constable, R. T. (2017) Using connectome-based predictive modeling to predict individual behavior from brain connectivity. Nature Protocols 12, 506–518.

Siless, V., Hubbard, N. A., Jones, R., Wang, J., Lo, N., Bauer, C. C. C., Goncalves, M., Frosch, I., Norton, D., Vergara, G., Conroy, K., De Souza, F. V., Rosso, I. M., Wickham, A. H., Cosby, E. A., Pinaire, M., Hirshfeld-Becker, D., Pizzagalli, D. A., Henin, A., Hofmann, S. G., Auerbach, R. P., Ghosh, S., Gabrieli, J., Whitfield-Gabrieli, S. and Yendiki, A. (2020) Image acquisition and quality assurance in the Boston Adolescent Neuroimaging of Depression and Anxiety study. NeuroImage. Clinical 26, 102242.

Smith, S. M., Beckmann, C. F., Andersson, J., Auerbach, E. J., Bijsterbosch, J., Douaud, G., Duff, E., Feinberg, D. A., Griffanti, L., Harms, M. P., Kelly, M., Laumann, T., Miller, K. L., Moeller, S., Petersen, S., Power, J., Salimi-Khorshidi, G., Snyder, A. Z., Vu, A. T., Woolrich, M. W., Xu, J., Yacoub, E., Ugurbil, K., Van Essen, D. C., Glasser, M. F. and Consortium, W. U.-M. H. (2013) Resting-state fMRI in the Human Connectome Project. NeuroImage 80, 144–168.

Solmi, M., Radua, J., Olivola, M., Croce, E., Soardo, L., Salazar de Pablo, G., Il Shin, J., Kirkbride, J. B., Jones, P., Kim, J. H., Kim, J. Y., Carvalho, A. F., Seeman, M. V., Correll, C. U. and Fusar-Poli, P. (2022) Age at onset of mental disorders worldwide: large-scale meta-analysis of 192 epidemiological studies. Molecular psychiatry 27, 281–295.

Tebaldi, C. and Knutti, R. (2007) The use of the multi-model ensemble in probabilistic climate projections. Philos Trans A Math Phys Eng Sci 365, 2053–2075.

